# Trajectories of compliance with COVID-19 related guidelines: longitudinal analyses of 50,000 UK adults

**DOI:** 10.1101/2021.04.13.21255336

**Authors:** Liam Wright, Andrew Steptoe, Daisy Fancourt

## Abstract

**Background:** Governments have implemented a range of measure to tackle COVID-19, primarily focusing on changing citizens’ behaviours in order to lower transmission of the virus. Some policymakers have expressed concern that citizens would not maintain high levels of compliance with these behaviours over the pandemic and would instead exhibit so-called “behavioural fatigue”. While the concept has been criticized, there have been few tests of behavioural fatigue using data from the COVID-19 pandemic, and none that have tracked individuals’ compliance trajectories.

**Methods:** We used longitudinal data on self-reported compliance from 50,851 adults in the COVID-19 Social Study collected across two waves of the pandemic in the UK (01 April 2020 – 22 February 2021). We modelled typical compliance trajectories using latent growth curve analysis (LGCA) and tested for behavioural fatigue by attempting to identify a set of participants whose compliance decreased substantially over the study period.

**Results:** We selected a four-class LGCA solution. Most individuals maintained high levels of compliance over the pandemic and reported similar levels of compliance across the first and second waves. Approximately one in seven participants had decreasing levels of compliance across the pandemic, reporting noticeably lower levels of compliance in the second wave, a pattern compatible with behavioural fatigue. Individuals with declining compliance levels differed from those with consistently high compliance on multiple characteristics, including (young) age, better physical health, lower empathy and conscientiousness and greater general willingness to take risks.

**Conclusion:** While a minority, not all individuals have maintained high compliance across the pandemic. Decreasing compliance is related to several psychological traits. The results suggest that targeting of behaviour change messages later in the pandemic may be needed to increase compliance.

## Introduction

Prior to the full roll-out of a vaccine, government strategies to reduce the spread of COVID-19 have focused on changing citizens’ behaviours, for instance via advertising personal hygiene reminders (e.g., washing hands), mandating the wearing of face masks, recommending social distancing in public spaces, and prohibiting household mixing. Where followed, these interventions can reduce the spread of the virus (Chu et al., 2020). However, each require voluntary cooperation on behalf of citizens, sometimes at considerable personal cost. Compliance with these behaviours is high but not complete (Ipsos MORI, 2021; YouGov, 2021). International data shows that average levels of compliance have declined since the start of the pandemic, though compliance has increased somewhat as countries have experienced second waves (YouGov, 2021; Petherick et al., 2021).

While population-level trends have been mapped, these trends could mask considerable heterogeneity: some individuals may have maintained high levels of compliance, while others may have stopped. Existing evidence from the current and previous pandemics shows that trends in compliance can differ markedly across groups (Jørgensen et al., 2021; Petherick et al., 2021; van der Weerd et al., 2011; Wright & Fancourt, 2020). For instance, in the UK, compliance decreased faster among younger age groups over the first five months of the COVID-19 pandemic in the UK (Wright & Fancourt, 2020). Yet, to our knowledge, no research has been carried out looking at individual compliance trajectories across the COVID-19 pandemic. This is a striking gap given that variation in infectiousness can influence how viruses spread (Bansal et al., 2007; Lloyd-Smith et al., 2005) and that examining individual compliance trajectories could support the targeting and design of interventions for behaviour change.

Modelling compliance trajectories is also important for understanding the extent to which individuals have suffered “behavioural fatigue”, understood here as a loss of motivation to comply as pandemic progress, holding other things (such as background risk of infection) constant (Petherick et al., 2021; WHO Regional Office for Europe, 2020). At the beginning of the pandemic, lead UK Government scientists cited behavioural fatigue as a reason to delay the imposition of strict lockdown (Mahase, 2020). The concept was widely criticised as being poorly elucidated and lacking scientific basis (Abbasi, 2020; Drury et al., 2021; Hahn et al., 2020; Michie et al., 2020; Reicher & Drury, 2021; Sibony, 2020). Indeed, there continues to be disagreement on how behavioural fatigue should be defined (Bell, 2020; Harvey, 2020; Lilleholt et al., 2020; Michie et al., 2020; Petherick et al., 2021), and related phrases have entered also usage, notably “pandemic fatigue” (Petherick et al., 2021; WHO Regional Office for Europe, 2020).

Despite these criticisms – and the apparent importance of the idea in influencing policy – empirical testing of behavioural fatigue has been limited. Recent work has studied declines in population-level compliance (Petherick et al., 2021), focused on between-person differences in compliance motivations (Lilleholt et al., 2020), or assessed the role of related factors, such as boredom, in predicting compliance behaviour (Martarelli & Wolff, 2020). While informative, these approaches do not evaluate changes in compliance at the level of the individual, a process central to the notion of fatigue (Harvey, 2020; Michie et al., 2020). Studying average compliance levels offers only a blunt test of behavioural fatigue as averages may mask significant heterogeneity.

In this study, we used (unbalanced) panel data from 50,000 adults from across two waves of the COVID-19 pandemic in the UK (April 2020 – February 2021) to model individual trajectories of (self-reported) compliance with COVID-19 guidelines. We used latent class growth analysis to identify “typical” compliance trajectories (Herle et al., 2020), examined whether these were compatible with behavioural fatigue, and tested how compliance trajectories were related to a variety of demographic, personality trait and individual risk factors. Our test of behavioural fatigue was whether we could identify a sizeable proportion of participants whose compliance decreased substantially over the study period. Note, while such a pattern is compatible with behavioural fatigue, it is not conclusive evidence thereof: though death rates were greater at the height of the second wave and similar lockdown measure were been put in place, many relevant factors relevant to compliance changed, including understanding of rules (Denford et al., 2020; Williams et al., 2020; Williams & Dienes, 2021), information seeking (though this may be a consequence of fatigue; Lilleholt et al., 2020), and scientific knowledge of the virus (Smith et al., 2020). Nevertheless, the study represents the first test of the consequences of behavioural fatigue at an individual level.

## Methods

### Participants

We used data from the COVID-19 Social Study; a large panel study of the psychological and social experiences of over 70,000 adults (aged 18+) in the UK during the COVID-19 pandemic. The study commenced on 21 March 2020 and involved online weekly (from August 2020, monthly) data collection across the pandemic in the UK. The study is not random and therefore is not representative of the UK population, but it does contain a heterogeneous sample. The sample was recruited using three primary approaches. First, convenience sampling was used, including promoting the study through existing networks and mailing lists (including large databases of adults who had previously consented to be involved in health research across the UK), print and digital media coverage, and social media. Second, more targeted recruitment was undertaken focusing on (i) individuals from a low-income background, (ii) individuals with no or few educational qualifications, and (iii) individuals who were unemployed. Third, the study was promoted via partnerships with third sector organisations to vulnerable groups, including adults with pre-existing mental health conditions, older adults, carers, and people experiencing domestic violence or abuse. The study was approved by the UCL Research Ethics Committee [12467/005] and all participants gave informed consent. The study protocol and user guide (which includes full details on recruitment, retention, data cleaning, weighting and sample demographics) are available at https://github.com/UCL-BSH/CSSUserGuide.

For these analyses, we used data from the eleven months between 01 April 2020 to 22 February 2021. To model non-linear changes in compliance trajectories we focused on individuals with compliance data from three or more data collections across the study period (n = 50,851). This sample represents 71.2% of those with data collection by 22 February 2021. Lockdown measures were first announced in the UK on 23 March 2020. The study period overlaps with two waves of COVID-19. Government guidelines and the severity of lockdown measures changed frequently across the study period. Supplementary Figure S1 shows 7-day COVID-19 caseloads and confirmed deaths, along with the Oxford Policy Tracker (Hale et al., 2020), a numerical summary of policy stringency, over the study period.

### Measures

#### Compliance with COVID-19 Guidelines

Compliance with guidelines was measured at each data collection using a single-item measure: “Are you following the recommendations from authorities to prevent spread of Covid-19?”. The item was measured on a seven-point Likert scale (1 = “Not at all”, 7 = “Very much so”), and analysed as a continuous variable.

#### Predictors of Compliance

We assess the role of several predictors of compliance. We included variables for demographic and socio-economic characteristics, social and pro-social factors, physical and mental health, and personality traits. We selected these variables using the COM-B framework of health behaviour (Michie et al., 2011). The COM-B model posits that behaviour is determined by subjective and objective capability, opportunity for action, and autonomic and reflective motivation.

For capability to comply, we included variables for locus of control, resilience, neighbourhood crowding, annual household income, educational level, diagnosed psychiatric condition, ethnicity and the Big-5 personality trait conscientiousness. For opportunity to comply, we included variables for country of residence, employment status, neighbourhood crowding, and availability of neighbourhood space. For motivation to comply, we include variables for long-term physical health conditions (0, 1, 2+), age (grouped), sex, self-isolation during first the first wave, remaining Big-5 personality traits (openness, extraversion, agreeableness, and neuroticism), (cognitive and emotional) empathy, neighbourhood social capital, attachment to neighbourhood, neighbourhood satisfaction, risk-taking behaviour, household overcrowding (1+ persons per room), living arrangement (alone, not alone without child, not alone with child), and mental health experiences during the first lockdown (same, better or worse vis-à-vis prior to the pandemic). Several of these variables were measured in one-off modules during follow-up and so are missing for many individuals. More detail on the individual measures is provided in the Supplementary Information. These variables have been studied previously in analyses of the COVID-19 Social Study (Wright, Steptoe, et al., 2021; Wright & Fancourt, 2020).

### Statistical Analysis

Our analysis proceeded in three steps. First, we estimated a growth curve model to examine between-person variation in compliance trajectories. To allow for compliance to change non-linearly with time, we modelled growth curves using natural splines (three degrees of freedom) and included random intercepts and random slopes in this model. Second, we used used latent class growth analysis (LCGA) to identify “typical” compliance trajectories (Herle et al., 2020), again using natural splines to allow for flexible relationships with time. We repeated LCGA models for 2-7 classes, using a thresholds link function to account for the non-normality of our compliance measure. We selected the final model considering the Bayes Information Criterion (BIC) and entropy values, average latent class probabilities and substantive interpretation of the classes identified. To reduce the risk of the algorithm identifying a local maximum, we fit models with 100 random starts (30 iterations each).

Third, we used multinomial logistic regression to identify predictors of class membership. For each variable, we first estimated a bivariate model and then estimated a multivariate model that included adjustment for sex, country, shielding, psychiatric diagnoses and long-term conditions, household overcrowding, living arrangement, income, (baseline) employment status, ethnic group, education, age group, and Big-5 personality traits (Soto & John, 2017). To account for uncertainty in the LCGA classes, we estimated multinomial regressions using “pseudo” draws from posterior probability matrix (Nylund-Gibson et al., 2019). We combined this procedure with multiple imputation (m = 60) to account for item missingness, pooling estimates using Rubin’s Rules (Rubin, 1987). We used unweighted data to estimate growth curve and LCGA models but added weights in multinomial models. The weights were created using entropy balancing according to population proportions for age, gender, ethnicity, education, and country of living (Nomis, 2018). The data used to create these weights was missing for 357 participants, so the sample is slightly smaller (0.7%) for the multinomial logit models (n = 50,494).

Data analysis was carried out in R v 4.0.3. (R Core Team, 2020). The growth curve model was estimated using the lme4 package (Bates et al., 2015), LCGA models were estimated using the lcmm package (Proust-Lima et al., 2017), multinomial regression was carried out the nnet package (Venables et al., 2002), and imputed data was generated using the mice package (van Buuren & Groothuis-Oudshoorn, 2011). Due to stipulations set out by the ethics committee, data will be made available at the end of the pandemic. The code to replicate the analysis is available at https://osf.io/hmn9s/.

### Role of the Funding Source

The funders had no final role in the study design; in the collection, analysis, and interpretation of data; in the writing of the report; or in the decision to submit the paper for publication. All researchers listed as authors are independent from the funders and all final decisions about the research were taken by the investigators and were unrestricted.

## Results

### Descriptive Statistics

Sample descriptive statistics are displayed in Table 1. Table S1 shows descriptive statistics by last month of (continuous) follow-up or whether the participant was ineligible for inclusion for the study. There is evidence of differences in attrition rates across groups. Notably older individuals were more likely to remain in the study. Figure S2 shows trends in compliance by last month of follow-up. Those with higher compliance levels were more likely to remain in the study, but there were qualitatively similar trends in compliance across groups: average compliance decreased from the first lockdown to early Autumn before increasing as the UK faced its second wave.

**Table 1:** Descriptive statistics. For continuous variables, Mean (SD). For categorical variables, n (%).

| Variable |  | Unweighted<br>Observed | Weighted<br>Imputed Data | Missing % |
| --- | --- | --- | --- | --- |
| n |  | 50,851 | 50,494 |  |
| Age (grouped) | 18-29 | 3,674 (7.23%) | 9,843.37<br>(19.49%) | 0% |
|  | 30-45 | 13,576 (26.7%) | 13,177.79<br>(26.1%) |  |
|  | 46-59 | 16,495<br>(32.44%) | 12,171.91<br>(24.11%) |  |
|  | 60+ | 17,106<br>(33.64%) | 15,300.92<br>(30.3%) |  |
| Gender | Male | 12,406 (24.5%) | 24,936.48<br>(49.39%) | 0.43% |
|  | Female | 38,228 (75.5%) | 25,557.52<br>(50.61%) |  |
| Ethnicity | White | 48,383<br>(95.46%) | 44,032.83<br>(87.2%) | 0.32% |

|  | Variable | Unweighted<br>Observed | Weighted<br>Imputed Data | Missing % |
| --- | --- | --- | --- | --- |
|  | Non-White | 2,303 (4.54%) | 6,461.17 (12.8%) |  |
| Country | England | 41,148<br>(80.92%) | 42,557.58<br>(84.28%) | 0% |
|  | Wales | 5,938 (11.68%) | 2,383.76 (4.72%) |  |
|  | Scotland | 3,239 (6.37%) | 4,139.31 (8.2%) |  |
|  | Northern Ireland | 526 (1.03%) | 1,413.35 (2.8%) |  |
| Education | GCSE or below | 6,995 (13.76%) | 16,501.88<br>(32.68%) | 0% |
|  | A-Level | 8,820 (17.34%) | 17,107.77<br>(33.88%) |  |
|  | Degree or above | 35,036 (68.9%) | 16,884.36<br>(33.44%) |  |
| Employment Status | Retired | 12,224<br>(24.04%) | 10,780.01<br>(21.35%) | 0% |
|  | Employed | 31,663<br>(62.27%) | 28,525.58<br>(56.49%) |  |
|  | Student | 1,599 (3.14%) | 4,500.90 (8.91%) |  |
|  | Unemployed/Inactive | 5,365 (10.55%) | 6,687.51<br>(13.24%) |  |
| Household Income | < £16k | 6,695 (14.54%) | 10,445.43<br>(20.69%) | 9.45% |
|  | £16k - £30k | 11,133<br>(24.18%) | 13,797.00<br>(27.32%) |  |
|  | £30k - £60k | 16,186<br>(35.15%) | 16,287.24<br>(32.26%) |  |
|  | £60k - £90k | 7,131 (15.49%) | 6,082.37<br>(12.05%) |  |
|  | £90k+ | 4,899 (10.64%) | 3,881.95 (7.69%) |  |
| Living Arrangement | Not alone, no child | 27,867 (54.8%) | 28,695.19<br>(56.83%) | 0% |
|  | Not alone, with child | 13,046<br>(25.66%) | 12,762.13<br>(25.27%) |  |
|  | Alone | 9,938 (19.54%) | 9,036.69 (17.9%) |  |
| Overcrowding | <1 persons per room | 45,700<br>(89.87%) | 42,293.91<br>(83.76%) | 0% |
|  | 1+ person per room | 5,151 (10.13%) | 8,200.09<br>(16.24%) |  |
| Lockdown 1.0 Mental Health | Same | 17,236<br>(59.32%) | 28,709.91<br>(56.86%) | 42.86% |
|  | Worse | 9,543 (32.84%) | 17,935.09<br>(35.52%) |  |
|  | Better | 2,277 (7.84%) | 3,849.00 (7.62%) |  |
| Shielding (pre-existing<br>condition) | No | 42,673<br>(92.91%) | 46,951.19<br>(92.98%) | 9.68% |
|  | Yes | 3,257 (7.09%) | 3,542.81 (7.02%) |  |
| Psychiatric condition | No | 41,522<br>(81.65%) | 40,186.76<br>(79.59%) | 0% |
|  | Yes | 9,329 (18.35%) | 10,307.24<br>(20.41%) |  |
| Long-Term Conditions | 0 | 28,079<br>(58.42%) | 29,362.49<br>(58.15%) | 5.48% |
|  | 1 | 12,845<br>(26.72%) | 13,088.76<br>(25.92%) |  |
|  | 2+ | 7,141 (14.86%) | 8,042.75<br>(15.93%) |  |
|  | Openness | 15.4 (3.29) | 14.88 (3.34) | 0% |
|  | Conscientiousness | 15.93 (2.97) | 15.56 (3.1) | 0% |
|  | Extraversion | 12.91 (4.29) | 12.55 (4.32) | 0% |

| Variable | Unweighted<br>Observed | Weighted<br>Imputed Data | Missing % |
| --- | --- | --- | --- |
| Agreeableness | 15.57 (3.06) | 15.35 (3.16) | 0% |
| Neuroticism | 11.32 (4.32) | 11.58 (4.51) | 0% |
| Resilience | 20.2 (5.17) | 19.85 (5.37) | 32.42% |
| Optimism | 19.76 (4.7) | 18.73 (4.85) | 39.62% |
| (External) Locus of control | 12.26 (2.64) | 12.73 (2.76) | 39.73% |
| Risk-taking | 4.39 (2.35) | 4.41 (2.4) | 50.47% |
| Cognitive Empathy | 18.72 (4.83) | 18.03 (4.96) | 41.83% |
| Emotional Empathy | 20.77 (4.64) | 20.07 (4.84) | 42.08% |
| Neighbourhood Social<br>Capital | 16.95 (3.49) | 16.38 (3.67) | 48.18% |
| Neighbourhood Attachment | 10.86 (3.25) | 10.2 (3.43) | 47.83% |
| Neighbourhood Satisfaction | 4.08 (0.93) | 3.94 (0.98) | 47.58% |
| Neighbourhood Space | 8.4 (1.15) | 8.26 (1.26) | 48.41% |
| (Low) Neighbourhood<br>Crowding | 6.97 (1.87) | 6.9 (1.89) | 48.24% |

### Growth Curve Modelling

Results from the growth curve model are displayed in Figure 1. The plot shows trends in the 2.5^th^, 50^th^ and 97.5^th^ percentiles of predicted compliance values over the study period. Figure S3 plots predicted compliance trends for a subsample of 6,000 participants in the study. The qualitative trends of declining and then increasing compliance levels is displayed among most individuals but is pronounced among a set of individuals whose compliance decreases substantially. Among those who compliance declined the most, there was less pronounced increases in compliance during the second wave. For individuals whose compliance decreased to a lesser extent, compliance returned to broadly similar levels as reported in the first levels. The plots demonstrate that population-level trends mask substantial heterogeneity.

**Figure 1:**
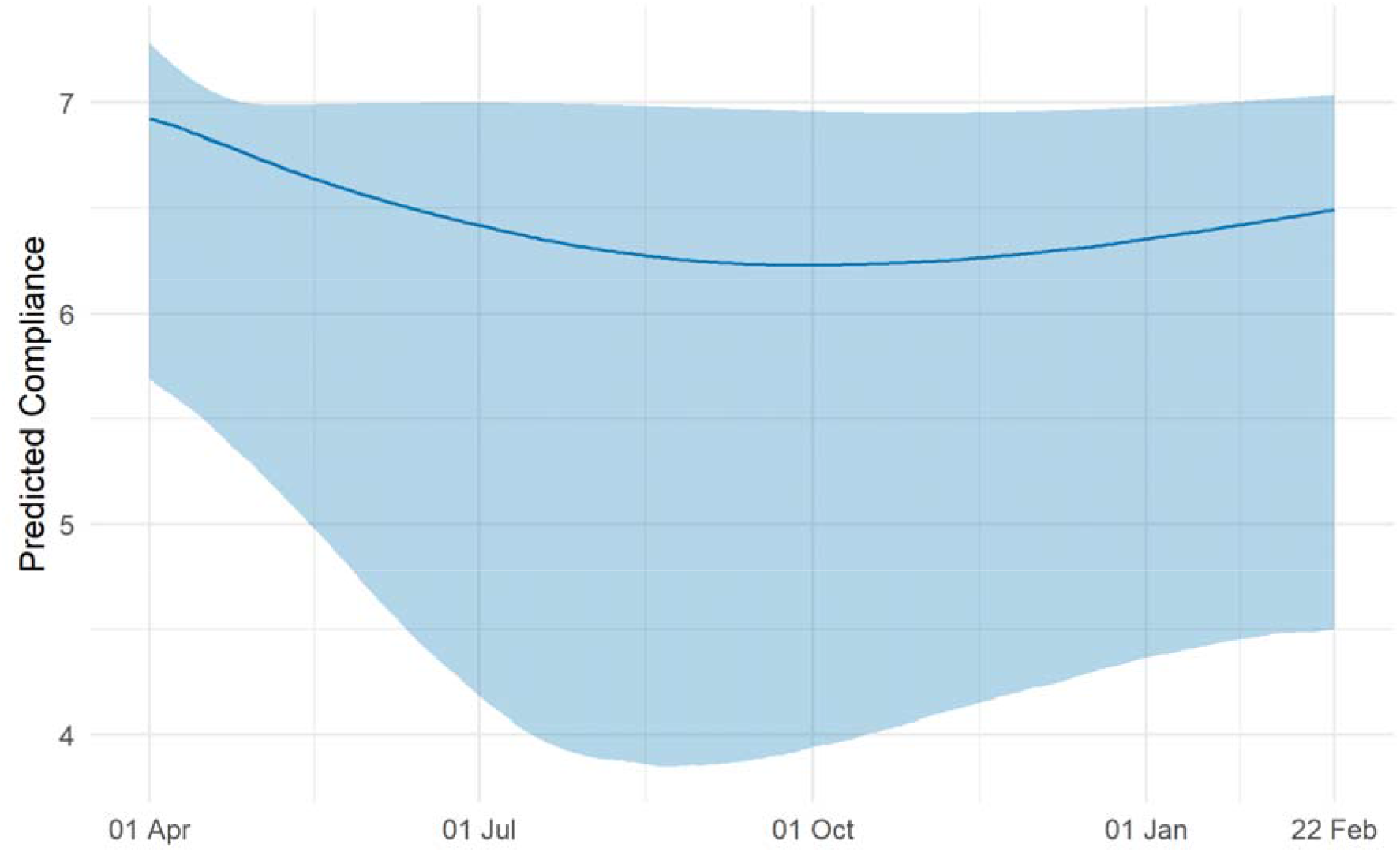
Trends in median compliance level derived from growth curve model with time modelled with natural splines with degrees of freedom 3. Bands represent trends in 2.5^th^ and 97.5^th^ percentiles of predicted compliance levels.

### Compliance Trajectory Classes

Fit statistics from the LGTA models are displayed in Figure S4. We selected a 4-class solution (entropy = 0.82) as solutions with a higher number of classes yielded substantively similar groups. Table 2 displays class proportions and average class probabilities. Figure 2 displays predicted compliance trends in each group. Results are alternatively displayed as predicted probabilities in Figure S5 and as a sample of growth curves modelled in the previous section in Figure S6. The largest group (Class 1; 32.8% of weighted observations) consisted of individuals whose compliance remained high throughout the pandemic. Classes 2 and 3 (28.7% and 24%, respectively) consisted of individuals whose compliance began high before dropping across summer and increasing to approximately its former levels during the second wave. Class 4 (14.6%), on the other hand, consisted of individuals whose compliance decreased sharply over the first lockdown and, while rising during the second wave, did not reach its former levels. This pattern is consistent with behavioural fatigue, though it should be noted predicted levels of compliance during February 2020 were still predicted to be approximately 5 on a 1-7 scale.

**Table 2:** Class proportions and class probabilities by most likely class, derived from four-class LGCA model.

| Most Likely Class | Class Proportions |  | Average Class Probability |  |  |  |
| --- | --- | --- | --- | --- | --- | --- |
|  | Unweighted | Weighted | Class 1 | Class 2 | Class 3 | Class 4 |
| Class 1 | 34.6% | 32.8% | 0.089 | 0.906 | 0.005 | 0.000 |
| Class 2 | 28.1% | 28.7% | 0.063 | 0.004 | 0.889 | 0.044 |
| Class 3 | 26.2% | 24% | 0.865 | 0.063 | 0.072 | 0.000 |
| Class 4 | 11% | 14.6% | 0.000 | 0.000 | 0.077 | 0.923 |

**Figure 2:**
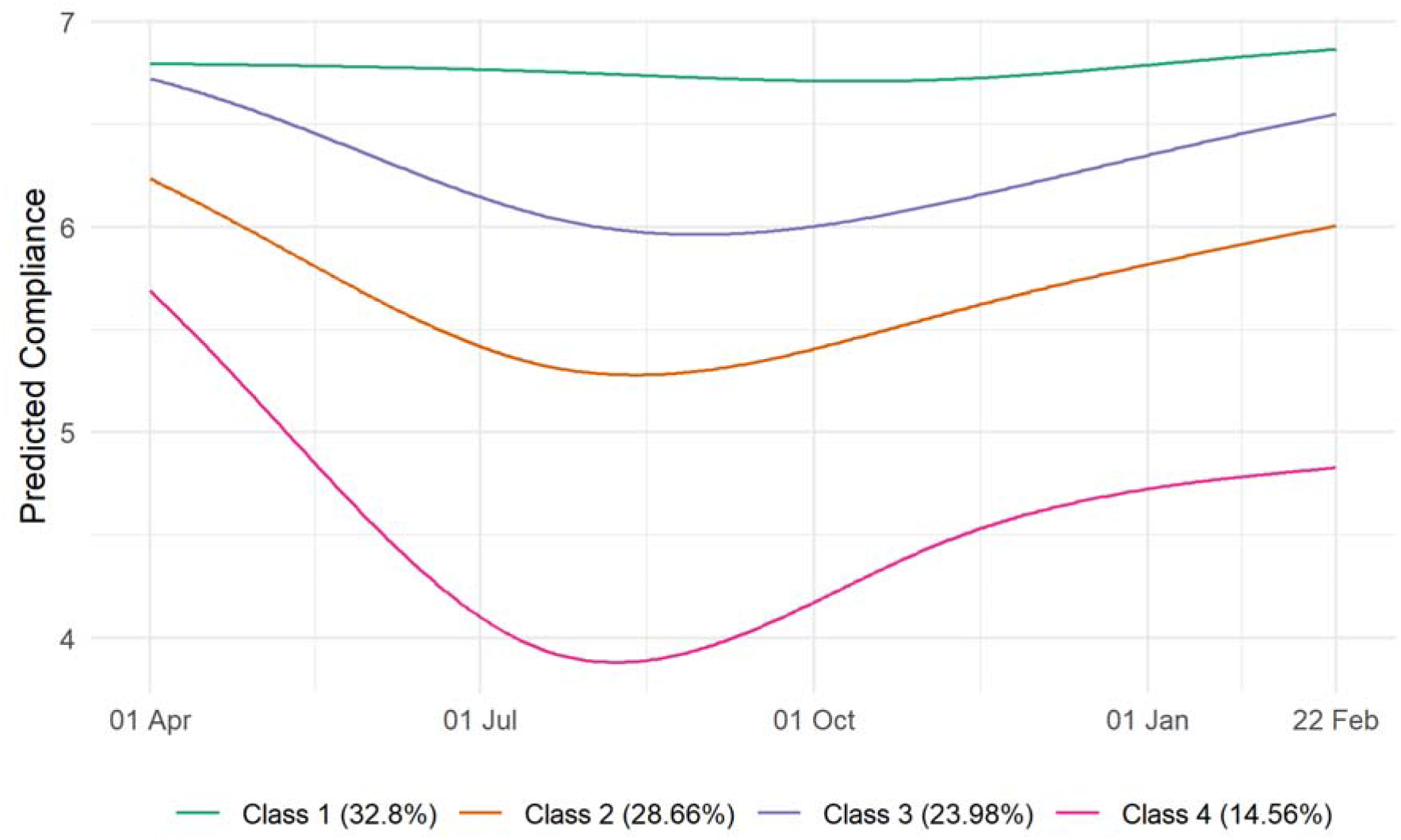
Predicted compliance trajectories by class, four-class LGCA.

### Predictors of Compliance Trajectories

The results of multivariate multinomial logistic regressions exploring the predictors of class membership are displayed in Figure 3 (personality traits) and Figure 4 (demographic, socioeconomic, health, and neighbourhood characteristics). (Bivariate regressions are displayed in Figures S7-S8.) For comparability, continuous variables are scaled such that a one unit change is equal to a differences of 2 SD (Gelman, 2008). Many of the variables were related to compliance trajectories, withs several showing strong associations with behavioural fatigue (Class 4), including risk-taking behaviour, young age, non-retired employment status, (low) emotional empathy and conscientiousness, and shielding due to personal health risk during the first lockdown.

**Figure 3:**
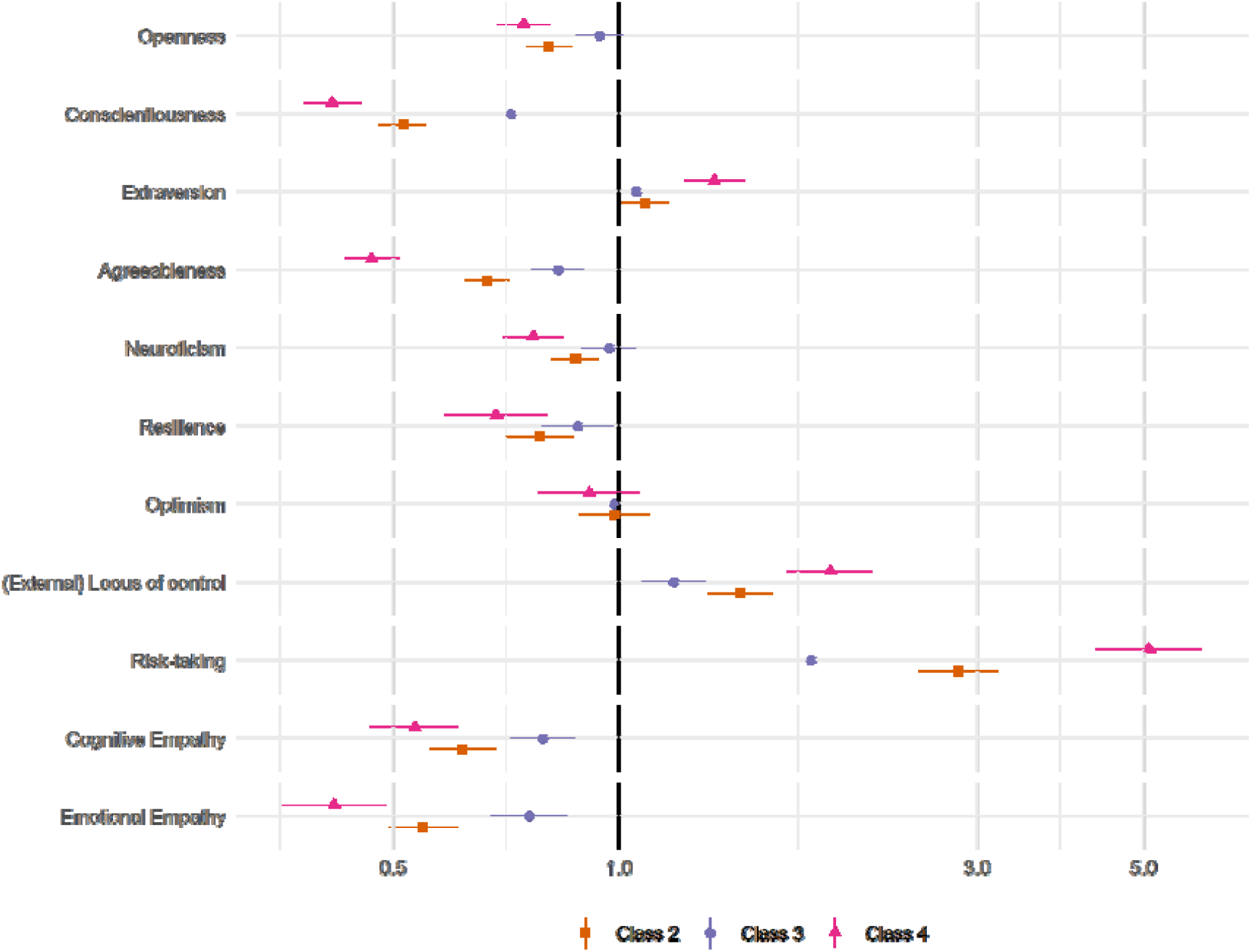
Results of multinomial logistic regression regressing (pseudo-)class membership on personality traits (reference class: Class 1). Adjustment for sex, country, shielding, psychiatric diagnoses and long-term conditions, household overcrowding, living arrangement, income, ethnic group, education, employment status, age group, and Big-5 personality traits. Models use weighted imputed data. Results pooled using Rubin’s (1987) rules.

**Figure 4:**
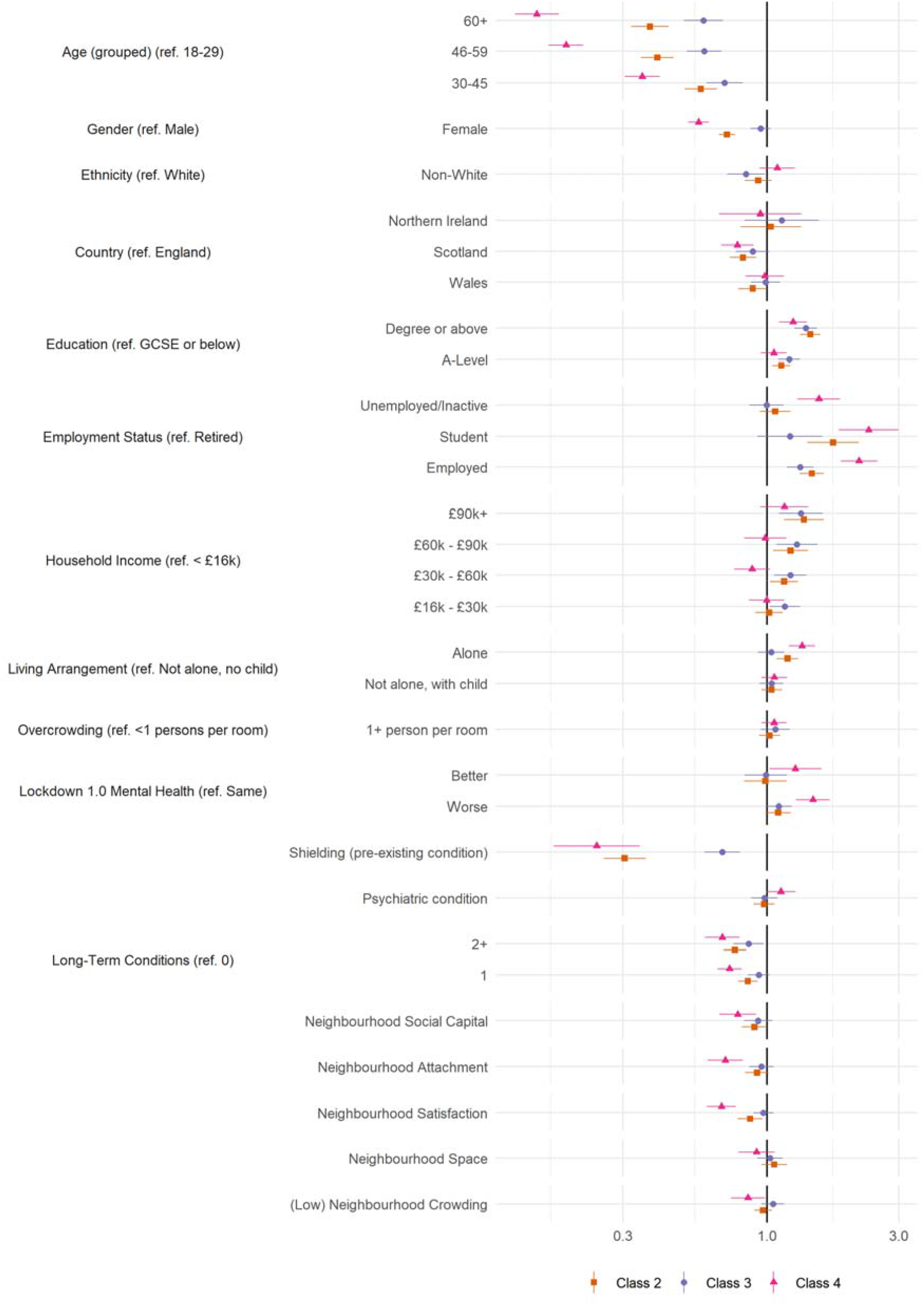
Results of multinomial logistic regression regressing (pseudo-)class membership on demographic, socioeconomic, health and neighbourhood factors (reference class: Class 1). Adjustment for sex, country, shielding, psychiatric diagnoses and long-term conditions, household overcrowding, living arrangement, income, ethnic group, education, employment status, age group, and Big-5 personality traits. Models use weighted imputed data. Results pooled using Rubin’s (1987) rules.

Also related to behavioural fatigue were (low) resilience, external locus of control, gender (male), low attachment or satisfaction with neighbourhood, and trait (low) neuroticism, (low) agreeableness, extraversion and openness to experience. Notably, associations between neighbourhood crowding, household overcrowding, or available space and compliance were small, and, relative to stable mental health, both improving *and* worsening mental health during first lockdown were related to lower compliance.

## Discussion

Using self-report data on compliance with COVID-19 guidelines from eleven month of the pandemic, we found evidence of substantial heterogeneity in compliance trajectories. While average compliance levels decreased only slightly from the first lockdown to mid-Autumn and returned to similar levels during the second wave in Winter 2020/2021, a minority of individuals for whom compliance decreased more substantially and did not fully recover, while the modal response was consistent high compliance levels. The results were consistent with at least some individuals experiencing behavioural fatigue. We identified several predictors of compliance trajectories. Notably, the behavioural fatigue pattern was related to age, (baseline) employment status, better physical health, and traits such as risk-taking behaviour, low empathy, low conscientiousness, disagreeableness, and low resilience.

The results are consistent with previous work on behavioural fatigue showing declines in average compliance across the pandemic (Petherick et al., 2021), though add richness in showing that it is only a minority of individuals for whom compliance decreases. The results suggest that low and decreasing compliance partly reflects risk (proxied by age and physical health) and personality traits. Some authors have argued that low compliance is largely a matter of material difficulties (Reicher & Drury, 2021), but this is not consistent with results here nor is it consistent with other studies that have identified several traits – including anti-social dark triad traits – as predictors of compliance (Blagov, 2020; Nowak et al., 2020; Zajenkowski et al., 2020).

While the results are compatible with behaviour fatigue, they are not dispositive. An issue is that several factors have changed across the pandemic that may also explain results. One alternative explanation for reduced compliance is “alert fatigue” (Williams & Dienes, 2021). Qualitative studies report that individuals have found it difficult to follow frequently-changing government rules (Denford et al., 2020; Williams et al., 2020; Wright, Burton, et al., 2021), leading to inadvertent non-compliance as well as bending of rules. However, during national lockdown in early 2021, rules were simplified and made largely uniform. Further, it is difficult to understand the role of traits, such as risk-taking, in predicting decreasing compliance in interpreting the results as alert fatigue. Other explanations are also possible, notably changes in perceptions of risk. While we are unable to assess this directly, we note that death rates were higher in the second wave (Figure S1). However, subjective risk perception may have reduced regardless, particularly for individuals who believed they had had the virus already. Disentangling changes in risk perception from behavioural fatigue is complicated by the possibility that reduced information seeking may be a consequence of fatigue (Lilleholt et al., 2020) and that individuals could employ motivated reasoning when willingness to comply has fallen (Rothgerber et al., 2020; Sembada & Kalantari, 2020; Shefrin, 2020). Regardless of the underlying cause, our results show heterogeneity in the level and trajectory of compliance behaviour. This has implications for transmission modelling, and for the targeting or design of interventions at those who have lowest compliance.

While we find that average compliance declined, it should be reiterated that sustained declines were a minority response. The majority of individuals reported high levels of compliance throughout the pandemic and reported similar levels of compliance in the first and second waves. However, three caveats should be noted. First, we used data from a convenience sample of individuals willing to participate – and continue participating in – a study expressly about COVID-19. These individuals are likely to comply with COVID-19 guidelines more than the wider population, so the extent of non-compliance may be underestimated in this study. Second, we modelled compliance as changing continuously through time, but individuals could violate guidelines intermittently to combat fatigue (for instance, occasionally meeting friends). Designs such as qualitative interviewing could be used to assess this possibility. Third, while our measure of compliance was framed in the present tense, it is possible that previous behaviour could influence responses, restricting temporal change. Nevertheless, our results are consistent with other research that has focused on specific behaviours (Petherick et al., 2021)

There were other limitations of our study. We used self-report compliance data, which is likely to be subject to issues of social desirability and recall bias. Some of the associations observed in the multinomial logistic regression modelling may be explained by non-differential measurement error. Attrition from the study meant that extrapolations further into the pandemic were made for many participants. Though, as noted, we suspect this means our estimates of non-compliance are conservative. Finally, we were unable to provide a conclusive test of behavioural fatigue. Innovative designs are required to separate fatigue from other alternative explanations.

Nevertheless, this study also had a number of strengths. To our knowledge, this is the first study investigating individual compliance trajectories. The results in this study may have implications for the modelling of the transmission of the virus, as well as raising questions for the targeting and design of behaviour change interventions. This study adds to a small literature examining compliance across the current and previous pandemics, showing variations in behaviour across time and between groups (Ayre et al., 2021; Cowling et al., 2010; Folmer, Brownlee, et al., 2020; Folmer, Kuiper, et al., 2020; Jørgensen et al., 2021; Petherick et al., 2021; Schneider et al., 2021; van der Weerd et al., 2011; Wright & Fancourt, 2020).

## Data Availability

Data used in this study will be made publicly available once the pandemic is over. The code used to run the analysis is available at https://osf.io/hmn9s/.

## Statements

### Declaration of interest

All authors declare no conflicts of interest.

### Funding

This Covid-19 Social Study was funded by the Nuffield Foundation [WEL/FR-000022583], but the views expressed are those of the authors and not necessarily the Foundation. The study was also supported by the MARCH Mental Health Network funded by the Cross-Disciplinary Mental Health Network Plus initiative supported by UK Research and Innovation [ES/S002588/1], and by the Wellcome Trust [221400/Z/20/Z]. DF was funded by the Wellcome Trust [205407/Z/16/Z]. The study was also supported by HealthWise Wales, the Health and Car Research Wales initiative, which is led by Cardiff University in collaboration with SAIL, Swansea University. The funders had no final role in the study design; in the collection, analysis and interpretation of data; in the writing of the report; or in the decision to submit the paper for publication. All researchers listed as authors are independent from the funders and all final decisions about the research were taken by the investigators and were unrestricted.

## Acknowledgements

The researchers are grateful for the support of a number of organisations with their recruitment efforts including: the UKRI Mental Health Networks, Find Out Now, UCL BioResource, HealthWise Wales, SEO Works, FieldworkHub, and Optimal Workshop.

## Author contributions

LW, AS and DF conceived and designed the study. LW analysed the data and wrote the first draft. All authors provided critical revisions. All authors read and approved the submitted manuscript.

## Notes

### Competing Interest Statement

The authors have declared no competing interest.

### Author Declarations

The study was approved by the UCL Research Ethics Committee [12467/005] and all participants gave informed consent.

### Summary of Updates

Due to an inconsistency between the data and the codebook, in an earlier version of this manuscript (posted April 15, 2021), the locus of control variable was incorrectly reverse coded. Results now correctly refer to the locus of control variable as increasing in external locus of control.

## References

Abbasi, K. (2020). Behavioural fatigue: A flawed idea central to a flawed pandemic response. BMJ, m3093. https://doi.org/10.1136/bmj.m3093

Ayre, J., Cvejic, E., McCaffery, K., Copp, T., Cornell, S., Dodd, R. H., Pickles, K., Batcup, C., Isautier, J. M. J., Nickel, B., Dakin, T., & Bonner, C. (2021). COVID-19 prevention behaviour over time in Australia: Patterns and long-term predictors from April to July 2020. MedRxiv, 2021.02.04.21251165. https://doi.org/10.1101/2021.02.04.21251165

Bansal, S., Grenfell, B. T., & Meyers, L. A. (2007). When individual behaviour matters: Homogeneous and network models in epidemiology. Journal of The Royal Society Interface, 4(16), 879–891. https://doi.org/10.1098/rsif.2007.1100

Bates, D., Mächler, M., Bolker, B., & Walker, S. (2015). Fitting Linear Mixed-Effects Models Using lme4. Journal of Statistical Software, 67(1). https://doi.org/10.18637/jss.v067.i01

Bell, V. (2020, March 20). Do we suffer ‘behavioural fatigue’ for pandemic prevention measures? Mind Hacks. https://mindhacks.com/2020/03/20/do-we-suffer-behavioural-fatigue-for-pandemic-prevention-measures/

Blagov, P. S. (2020). Adaptive and Dark Personality in the COVID-19 Pandemic: Predicting Health-Behavior Endorsement and the Appeal of Public-Health Messages. Social Psychological and Personality Science, 194855062093643. https://doi.org/10.1177/1948550620936439

Chu, D. K., Akl, E. A., Duda, S., Solo, K., Yaacoub, S., Schünemann, H. J., Chu, D. K., Akl, E. A., El-harakeh, A., Bognanni, A., Lotfi, T., Loeb, M., Hajizadeh, A., Bak, A., Izcovich, A., Cuello-Garcia, C. A., Chen, C., Harris, D. J., Borowiack, E., … Schünemann, H. J. (2020). Physical distancing, face masks, and eye protection to prevent person-to-person transmission of SARS-CoV-2 and COVID-19: A systematic review and meta-analysis. The Lancet, 395(10242), 1973–1987. https://doi.org/10.1016/S0140-6736(20)31142-9

Cowling, B. J., Ng, D. M. W., Ip, D. K. M., Liao, Q., Lam, W. W. T., Wu, J. T., Lau, J. T. F., Griffiths, S. M., & Fielding, R. (2010). Community Psychological and Behavioral Responses through the First Wave of the 2009 Influenza A(H1N1) Pandemic in Hong Kong. The Journal of Infectious Diseases, 202(6), 867–876. https://doi.org/10.1086/655811

Denford, S., Morton, K. S., Lambert, H., Zhang, J., Smith, L. E., Rubin, G. J., Cai, S., Zhang, T., Robin, C., Lasseter, G., Hickman, M., Oliver, I., & Yardley, L. (2020). Understanding patterns of adherence to COVID-19 mitigation measures: A qualitative interview study. MedRxiv, 2020.12.11.20247528. https://doi.org/10.1101/2020.12.11.20247528

Drury, J., Carter, H., Ntontis, E., & Guven, S. T. (2021). Public behaviour in response to the COVID-19 pandemic: Understanding the role of group processes. BJPsych Open, 7(1), e11. https://doi.org/10.1192/bjo.2020.139

Folmer, C. R., Brownlee, M., Fine, A., Kuiper, M. E., Olthuis, E., Kooistra, E. B., Bruijn, A. L. de, & Rooij, B. van. (2020). Social Distancing in America: Understanding Long-term Adherence to Covid-19 Mitigation Recommendations. PsyArXiv. https://doi.org/10.31234/osf.io/457em

Folmer, C. R., Kuiper, M., Olthuis, E., Kooistra, E. B., Bruijn, A. L. de, Brownlee, M., Fine, A., & Rooij, B. van. (2020). Maintaining Compliance when the Virus Returns: Understanding Adherence to Social Distancing Measures in the Netherlands in July 2020. PsyArXiv. https://doi.org/10.31234/osf.io/vx3mn

Gelman, A. (2008). Scaling regression inputs by dividing by two standard deviations. Statistics in Medicine, 27(15), 2865–2873. https://doi.org/10.1002/sim.3107

Hahn, U., Chater, N., Lagnado, D., Osman, M., & Raihani, N. (2020, March 16). Why a Group of Behavioural Scientists Penned an Open Letter to the U.K. Government Questioning Its Coronavirus Response. Behavioral Scientist. https://behavioralscientist.org/why-a-group-of-behavioural-scientists-penned-an-open-letter-to-the-uk-government-questioning-its-coronavirus-response-covid-19-social-distancing/

Hale, T., Angrist, N., Cameron-Blake, E., Hallas, L., Kira, B., Majumdar, S., Petherick, A., Phillips, T., Tatlow, H., & Webster, S. (2020). Oxford COVID-19 Government Response Tracker. Blavatnik School of Government. https://www.bsg.ox.ac.uk/research/research-projects/coronavirus-government-response-tracker

Harvey, N. (2020). Behavioral Fatigue: Real Phenomenon, Naïve Construct, or Policy Contrivance? Frontiers in Psychology, 11. https://doi.org/10.3389/fpsyg.2020.589892

Herle, M., Micali, N., Abdulkadir, M., Loos, R., Bryant-Waugh, R., Hübel, C., Bulik, C. M., & De Stavola, B. L. (2020). Identifying typical trajectories in longitudinal data: Modelling strategies and interpretations. European Journal of Epidemiology, 35(3), 205–222. https://doi.org/10.1007/s10654-020-00615-6

Ipsos MORI. (2021, January 17). Most Britons continue to say they are following coronavirus rules; almost half believe lockdown measures are not strict enough. https://www.ipsos.com/ipsos-mori/en-uk/most-britons-continue-say-they-are-following-coronavirus-rules-almost-half-believe-lockdown

Jørgensen, F., Bor, A., & Petersen, M. B. (2021). Compliance without fear: Individual[level protective behaviour during the first wave of the COVID□19 pandemic. British Journal of Health Psychology, bjhp.12519. https://doi.org/10.1111/bjhp.12519

Lilleholt, L., Zettler, I., Betsch, C., & Böhm, R. (2020). Pandemic Fatigue: Measurement, Correlates, and Consequences. PsyArXiv. https://doi.org/10.31234/osf.io/2xvbr

Lloyd-Smith, J. O., Schreiber, S. J., Kopp, P. E., & Getz, W. M. (2005). Superspreading and the effect of individual variation on disease emergence. Nature, 438(7066), 355–359. https://doi.org/10.1038/nature04153

Mahase, E. (2020). Covid-19: Was the decision to delay the UK’s lockdown over fears of “behavioural fatigue” based on evidence? BMJ, m3166. https://doi.org/10.1136/bmj.m3166

Martarelli, C. S., & Wolff, W. (2020). Too bored to bother? Boredom as a potential threat to the efficacy of pandemic containment measures. Humanities and Social Sciences Communications, 7(1), 1–5. https://doi.org/10.1057/s41599-020-0512-6

Michie, S., van Stralen, M. M., & West, R. (2011). The behaviour change wheel: A new method for characterising and designing behaviour change interventions. Implementation Science, 6(1), 42. https://doi.org/10.1186/1748-5908-6-42

Michie, S., West, R., & Harvey, N. (2020). The concept of “fatigue” in tackling covid-19. BMJ, m4171. https://doi.org/10.1136/bmj.m4171

Nomis. (2018). Annual Population Survey data. https://www.nomisweb.co.uk/

Nowak, B., Brzóska, P., Piotrowski, J., Sedikides, C., Żemojtel-Piotrowska, M., & Jonason, P. K. (2020). Adaptive and maladaptive behavior during the COVID-19 pandemic: The roles of Dark Triad traits, collective narcissism, and health beliefs. Personality and Individual Differences, 167, 110232. https://doi.org/10.1016/j.paid.2020.110232

Nylund-Gibson, K., Grimm, R. P., & Masyn, K. E. (2019). Prediction from Latent Classes: A Demonstration of Different Approaches to Include Distal Outcomes in Mixture Models. Structural Equation Modeling: A Multidisciplinary Journal, 26(6), 967–985. https://doi.org/10.1080/10705511.2019.1590146

Petherick, A., Goldszmidt, R., Andrade, E. B., Furst, R., Pott, A., & Wood, A. (2021). A Worldwide Assessment of COVID-19 Pandemic-Policy Fatigue (SSRN Scholarly Paper ID 3774252). Social Science Research Network. https://doi.org/10.2139/ssrn.3774252

Proust-Lima, C., Philipps, V., & Liquet, B. (2017). Estimation of Extended Mixed Models Using Latent Classes and Latent Processes: The R Package lcmm. Journal of Statistical Software, 78(2). https://doi.org/10.18637/jss.v078.i02

R Core Team. (2020). R: A language and environment for statistical computing (3.6.3) [Computer software]. R Foundation for Statistical Computing. https://www.R-project.org/

Reicher, S., & Drury, J. (2021). Pandemic fatigue? How adherence to covid-19 regulations has been misrepresented and why it matters. BMJ, 372, n137. https://doi.org/10.1136/bmj.n137

Rothgerber, H., Wilson, T., Whaley, D., Rosenfeld, D. L., Humphrey, M., Moore, A. L., & Bihl, A. (2020). Politicizing the COVID-19 Pandemic: Ideological Differences in Adherence to Social Distancing [Preprint]. PsyArXiv. https://doi.org/10.31234/osf.io/k23cv

Rubin, D. B. (1987). Multiple Imputation for Nonresponse in Surveys. John Wiley & Sons, Ltd. https://doi.org/10.1002/9780470316696

Schneider, C. R., Dryhurst, S., Kerr, J., Freeman, A. L. J., Recchia, G., Spiegelhalter, D., & Linden, S. van der. (2021). COVID-19 risk perception: A longitudinal analysis of its predictors and associations with health protective behaviours in the United Kingdom. Journal of Risk Research. https://www.tandfonline.com/doi/full/10.1080/13669877.2021.1890637

Sembada, A. Y., & Kalantari, H. D. (2020). Biting the travel bullet: A motivated reasoning perspective on traveling during a pandemic. Annals of Tourism Research, 103040. https://doi.org/10.1016/j.annals.2020.103040

Shefrin, H. (2020). The Psychology Underlying Biased Forecasts of COVID-19 Cases and Deaths in the United States. Frontiers in Psychology, 11, 590594. https://doi.org/10.3389/fpsyg.2020.590594

Sibony, A.-L. (2020). The UK COVID-19 Response: A Behavioural Irony? European Journal of Risk Regulation, 11(2), 350–357. https://doi.org/10.1017/err.2020.22

Smith, G. D., Blastland, M., & Munafò, M. (2020). Covid-19’s known unknowns. BMJ, 371, m3979. https://doi.org/10.1136/bmj.m3979

Soto, C. J., & John, O. P. (2017). The next Big Five Inventory (BFI-2): Developing and assessing a hierarchical model with 15 facets to enhance bandwidth, fidelity, and predictive power. Journal of Personality and Social Psychology, 113(1), 117–143. https://doi.org/10.1037/pspp0000096

van Buuren, S., & Groothuis-Oudshoorn, K. (2011). mice: Multivariate Imputation by Chained Equations in R. Journal of Statistical Software, 45(3). https://doi.org/10.18637/jss.v045.i03

van der Weerd, W., Timmermans, D. R., Beaujean, D. J., Oudhoff, J., & van Steenbergen, J. E. (2011). Monitoring the level of government trust, risk perception and intention of the general public to adopt protective measures during the influenza A (H1N1) pandemic in the Netherlands. BMC Public Health, 11(1), 575. https://doi.org/10.1186/1471-2458-11-575

Venables, W. N., Ripley, B. D., & Venables, W. N. (2002). Modern applied statistics with S (4th ed). Springer.

WHO Regional Office for Europe. (2020). Pandemic fatigue: Reinvigorating the public to prevent COVID-19: policy framework for supporting pandemic prevention and management (WHO/EURO:2020-1573-41324-56242; p. 28). World Health Organization. https://apps.who.int/iris/handle/10665/337574

Williams, S. N., Armitage, C. J., Tampe, T., & Dienes, K. (2020). Public perceptions and experiences of social distancing and social isolation during the COVID-19 pandemic: A UK-based focus group study. BMJ Open, 10(7), e039334. https://doi.org/10.1136/bmjopen-2020-039334

Williams, S. N., & Dienes, K. (2021, February 19). The public aren’t complacent, they’re confused— How the UK government created “alert fatigue”. The BMJ. https://blogs.bmj.com/bmj/2021/02/19/the-public-arent-complacent-they-are-confused-how-the-uk-government-has-created-alert-fatigue/

Wright, L., Burton, A., McKinlay, A., Steptoe, A., & Fancourt, D. (2021). Public Opinion about the UK Government during COVID-19 and Implications for Public Health: A Topic Modelling Analysis of Open-Ended Survey Response Data [Preprint]. Public and Global Health. https://doi.org/10.1101/2021.03.24.21254094

Wright, L., & Fancourt, D. (2020). Do predictors of adherence to pandemic guidelines change over time? A panel study of 21,000 UK adults during the COVID-19 pandemic [Preprint]. medRxiv. https://doi.org/10.1101/2020.11.10.20228403

Wright, L., Steptoe, A., & Fancourt, D. (2021). Patterns of compliance with COVID-19 preventive behaviours: A latent class analysis of 20,000 UK adults [Preprint]. Epidemiology. https://doi.org/10.1101/2021.03.16.21253717

YouGov. (2021). Personal measures taken to avoid COVID-19. https://yougov.co.uk/topics/international/articles-reports/2020/03/17/personal-measures-taken-avoid-covid-19

Zajenkowski, M., Jonason, P. K., Leniarska, M., & Kozakiewicz, Z. (2020). Who complies with the restrictions to reduce the spread of COVID-19?: Personality and perceptions of the COVID-19 situation. Personality and Individual Differences, 166, 110199. https://doi.org/10.1016/j.paid.2020.110199

