## Supplemental Information for "Trajectories of compliance with COVID-19 related guidelines: longitudinal analyses of 50,000 UK adults"

Supplementary Information

### Methods

#### Predictors of Compliance Behaviour

##### Demographics and Socio-Economic Position

We included the demographic variables for country of residence (England, Scotland, Wales, Northern Ireland), sex (male, female), ethnicity (White, Non-White) and age (18-29, 30-44, 45-59, 60+). We also included three variables for socio-economic position (SEP): annual income (< £16k, £16k - £30k, £30k - £60k, £60k - £90k, £90k +), education level (GCSE or below, A-levels or equivalent, degree or above), and employment status (retired, unemployed/inactive, student, employed). Each variable was measured at baseline interview.

##### Social and Prosocial Factors

We included three measures of pro-social motivations: cognitive empathy, emotional empathy, and social capital. Neighbourhood social capital before COVID-19 was measured between 09-16 July by combining four items from the social cohesion subscale of the Neighbourhood Scales (Mujahid et al., 2007) and a further item, “Before COVID-19, this was a close-knit neighbourhood”. Items were rated on a five-point scale (1 = “strongly disagree”, 5 = “strongly agree”), with items reflecting beliefs about trust, shared values, and willingness to help others. We used the sum score of items on social capital prior to COVID-19 (range 5 – 25). Higher values indicating higher social capital.

Empathy was assessed between 11-18 July using subscales for empathic concern from the Interpersonal Reactivity Index (IRI; Davis, 1983). (Subscales for fantasy and personal distress were not administered.) Empathic concern (also known as emotional empathy) consists of 7 items and captures feelings of warmth, concern, and compassion for others. Perspective-taking (also known as cognitive empathy) assesses efforts to adopt the perspectives of others (7 items). Items were rated on a five-point scale ranging (1 = “does not describe me well”, 5 = “describes me very well”). We used the sum Likert score, coding items such that higher scores indicate higher empathy (range 0 - 28).

We also assessed four aspects of individuals’ neighbourhoods, each measured between 09-16 July. Neighbourhood attachment was assessed with three five-point Likert items on whether the participant feels their neighbourhood is their home (1 = “just a place to live”, 5 = “home”), their attachment to their neighbourhood (1 = “no attachment”, 5 = “strong attachment”) and feelings of belonging in their community (1 = “don’t belong at all”, 5 = “belong strongly”). We used the sum Likert score (range 3-15) with higher scores indicating greater neighbourhood attachment.

Neighbourhood satisfaction was assessed with a five-point Likert item (1 = “very dissatisfied”, 5 = “very satisfied”). Neighbourhood space was measured using three items on satisfaction with neighbourhood walkability, usable green space, and presence of trees. Each was measure on a three-point scale (1 = “dissatisfied”, 2 = “neither satisfied nor dissatisfied”, 3 = “satisfied”), which we summed into a single score (range 3 – 9). Neighbourhood crowding was measured using three items on satisfaction with neighbourhood traffic density, noise, and levels of crowding. Each was measured on a three-point scale (categories as above), which we summed into a single score (range 3 – 9), such that higher scores indicated lower neighbourhood crowding.

Finally, we included two items on household overcrowding (< 1 person per room, 1+ persons per room), and living arrangement (alone, with adult but no child, with child). Both were measured at baseline interview.

##### Personality Traits

Personality was measured at baseline interview using the Big Five Inventory (BFI-2; Soto & John, 2017), which measures five domains and 15 facets: openness (intellectual curiosity, aesthetic sensitivity, and creative imagination), conscientiousness (organisation, productiveness, and responsibility), extraversion (sociability, assertiveness, and energy level), agreeableness (compassion, respectfulness, and trust) and neuroticism (anxiety, depression, and emotional volatility). Each item was scored on a 5-point scale (1 = “strongly disagree”, 5 = “strongly agree”). We use the sum Likert score for each domain (range 3 - 15). High values indicate high levels of the trait.

Resilience was assessed between 14 – 21 May with the 6-item Brief Resilience Scale (Smith et al., 2008), a widely used measure of individuals’ ability to recover from stress. Items are rated on a five-point scale (1 = “strongly disagree”, 5 = “strongly agree”). We used the sum Likert score, with items coded such that higher scores indicate higher resilience (range 6 - 30).

Optimism was collected between 04 – 11 June using the 10-item Life Orientation Test-Revised (LOT-R; Scheier et al., 1994). Items are rated on a five-point scale ranging from “strongly disagree” to “strongly agree”. We used the sum Likert score, coding items such that higher scores indicate greater optimism (range 6 - 30).

Locus of control was measured between 04 – 11 June using the 6-item Locus of Control Scale developed by the University of Washington Beyond High School Project (Hirschman & Almgren, 2012), and captures generalized expectancies about whether individuals can (internal) or cannot (external) control events and outcomes in their lives. Responses were rated on a four-point scale ranging from (1 = “strongly agree”, 4 = “strongly disagree”). We used the sum score of responses, with items coded such that higher values indicate more external locus of control.

Risk-taking was measured between 23-30 July with one item from the Dohmen Risk Taking Scale (Dohmen et al., 2011). Respondents rated the extent to which they generally see themselves as a person who is fully prepared to take risks, rated on an 11-point scale (0 = “not at all willing to take risks” to 10 = “very willing to take risks”).

##### Health and Confidence in Government

We included variables for long-term physical health conditions (0, 1, 2+) using a multiple-choice question on medical conditions. Included conditions were high blood pressure, diabetes, heart disease, lung disease, cancer, any other clinically-diagnosed chronic physical health conditions, or any disability. Psychiatric diagnosis (yes, no) was with the same multiple choice question using items on clinically diagnosed depression, clinically diagnosed anxiety, and any other clinically diagnosed mental health problem. The data was collected at baseline interview.

Mental health during first lockdown vis-à-vis prior to the pandemic was measured with a single item question (“How do you feel your mental health was affected during lockdown in April/May?”). The response categories were: my mental health got worse compared to before Covid-19; My mental health was about the same; my mental health got better compared to before Covid-19. This variable was included in the survey between 18-25 June.

### Figures

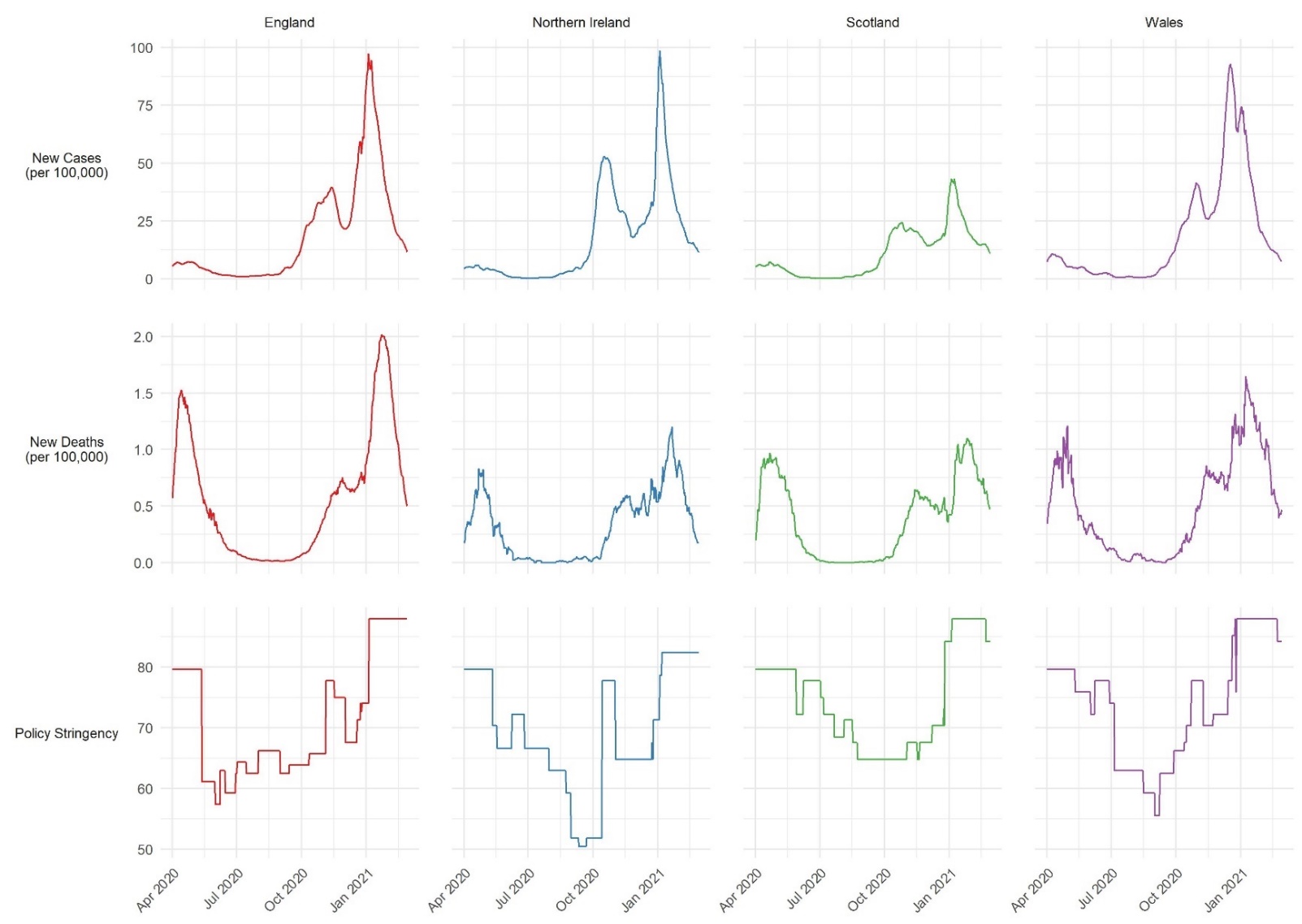

Figure S1: 7-day COVID-19 confirmed cases, COVID-19 deaths, and daily policy stringency by country. Source: Hale et al. (2020).

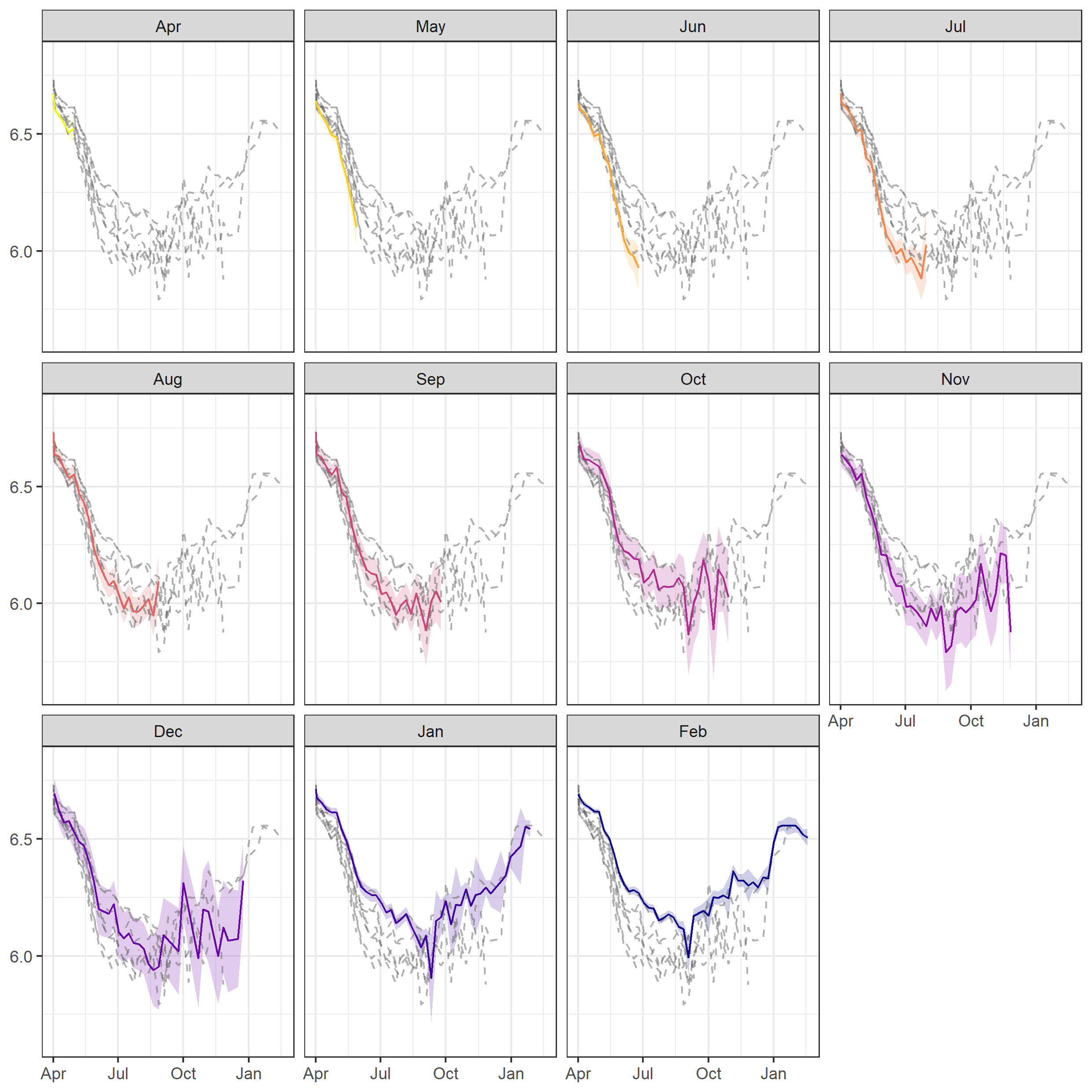

Figure S2: Trends in compliance by month of last interview. Dashed lines represent average compliance level by last month of interview. Coloured lines represent average compliance for the month listed in the panel title. Graphs show that individuals who remained in the study longer had higher compliance on average.

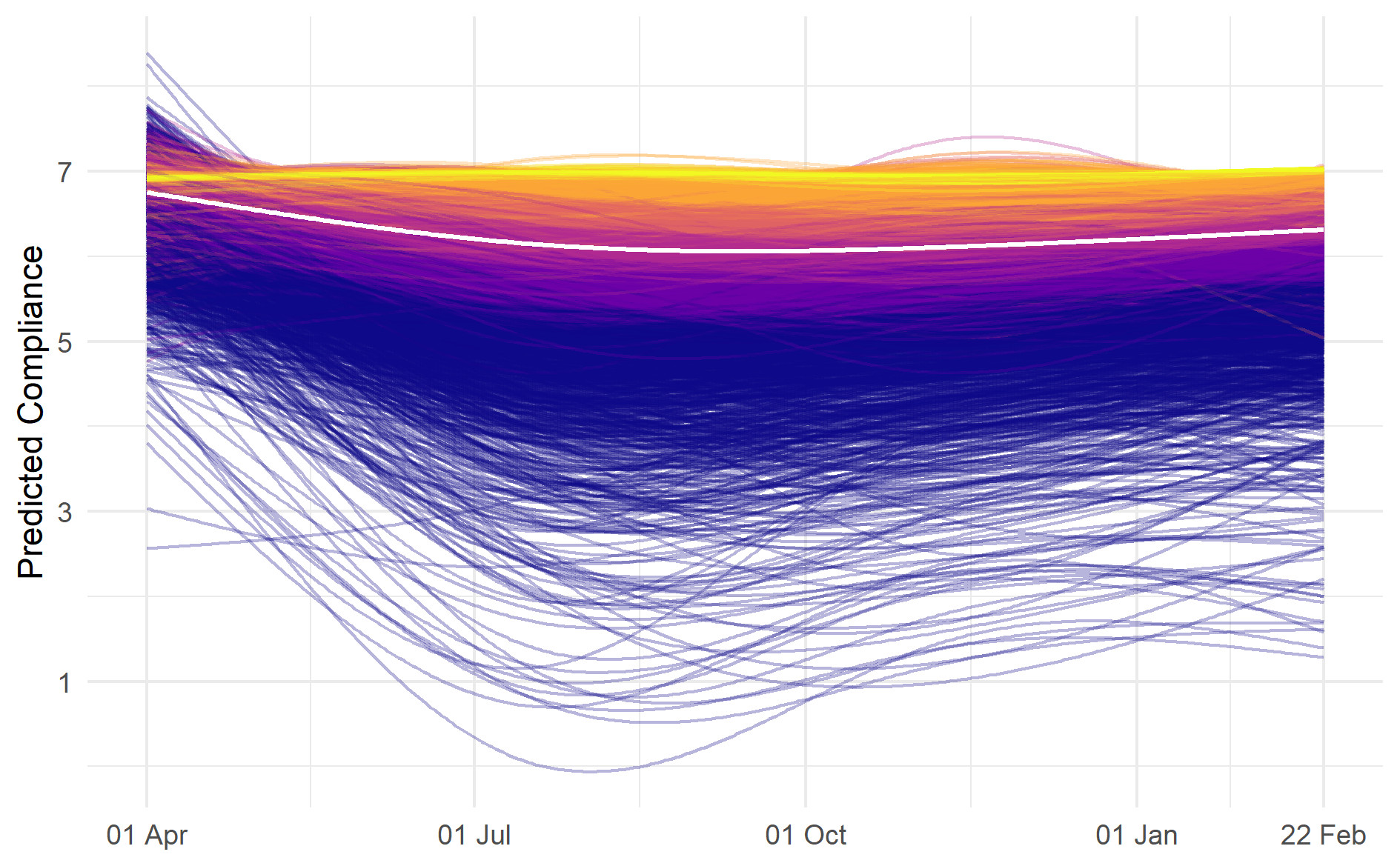

Figure S3: Predicted compliance trends for sample of 6,000 participants derived from growth curve models. Colors represent sextile of average compliance levels. Lighter colors indicate individuals with highest average level of compliance.

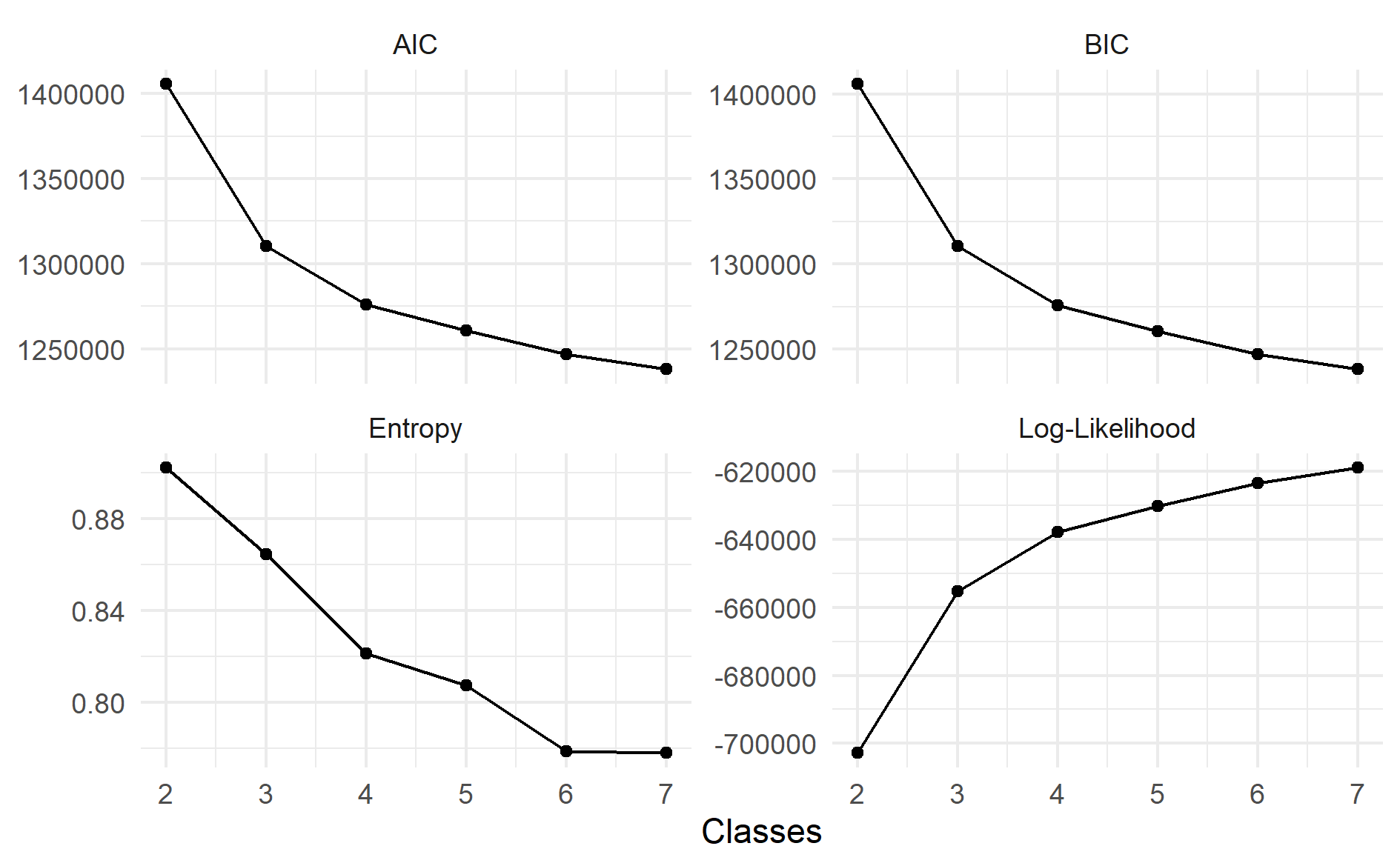

Figure S4: Fit statistics from LGCA models.

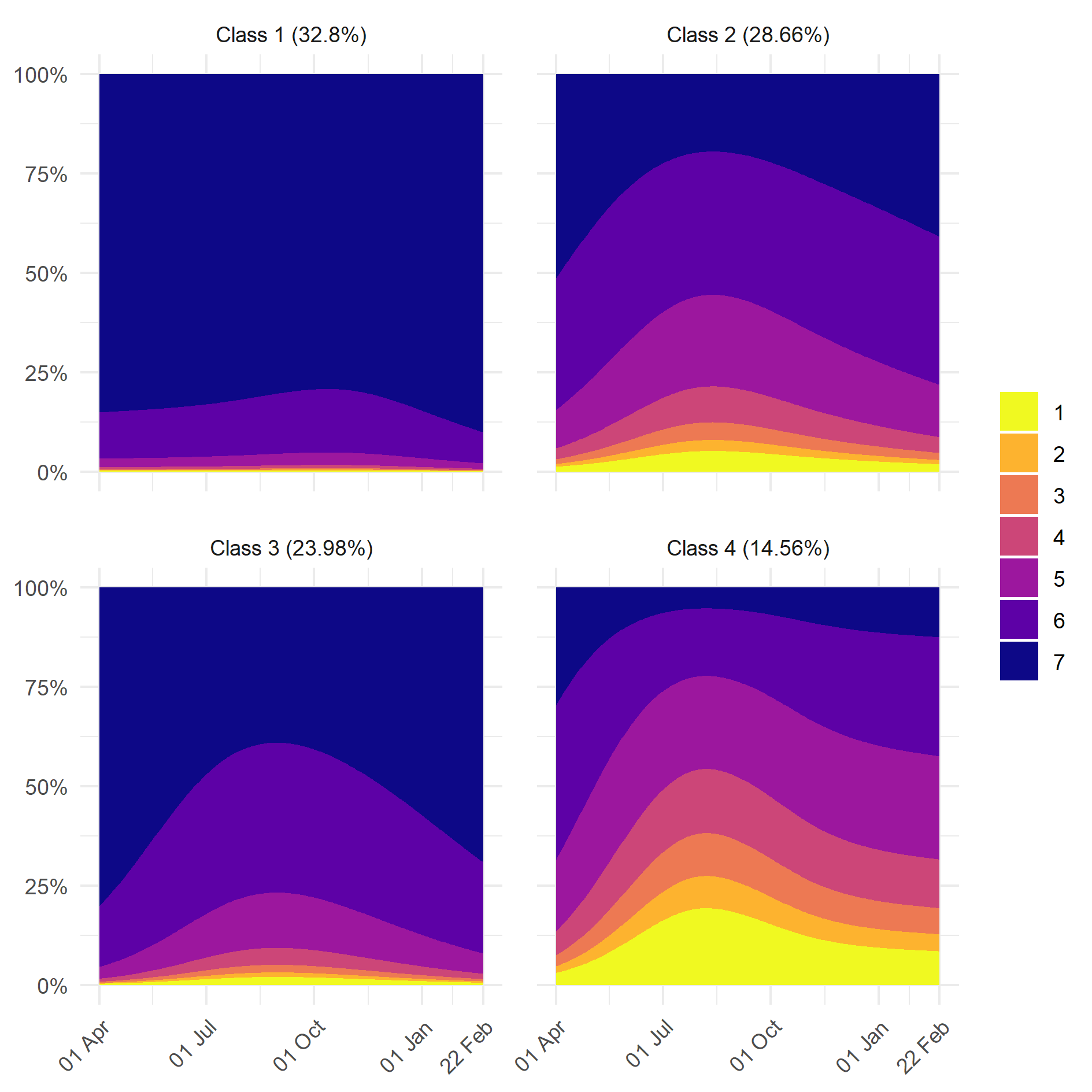

Figure S5: (Stacked) predicted probability of given compliance level by class and date.

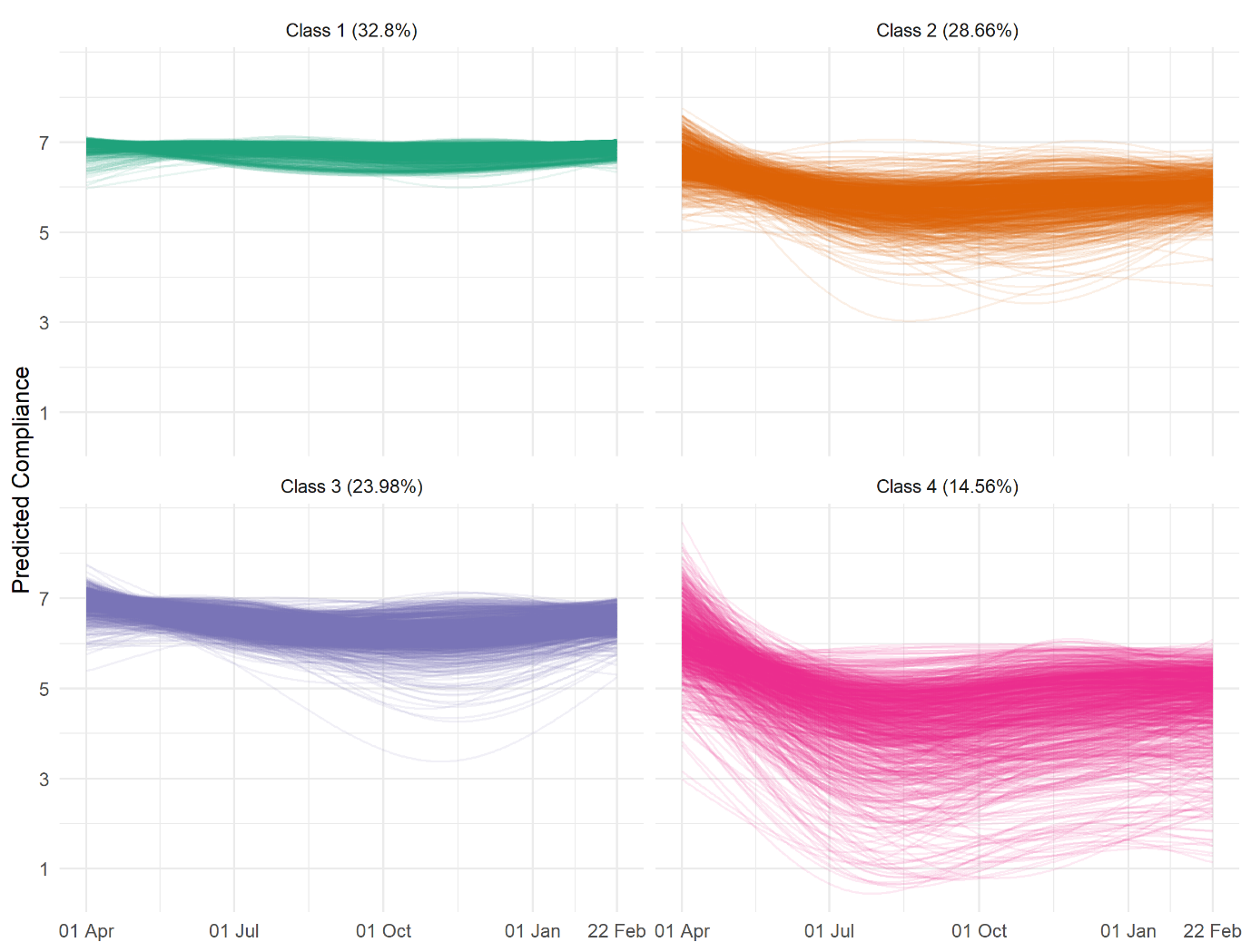

Figure S6: Random sample of predicting compliance trajectories derived from growth curve models by most-likely class membership from four-class LGCA model.

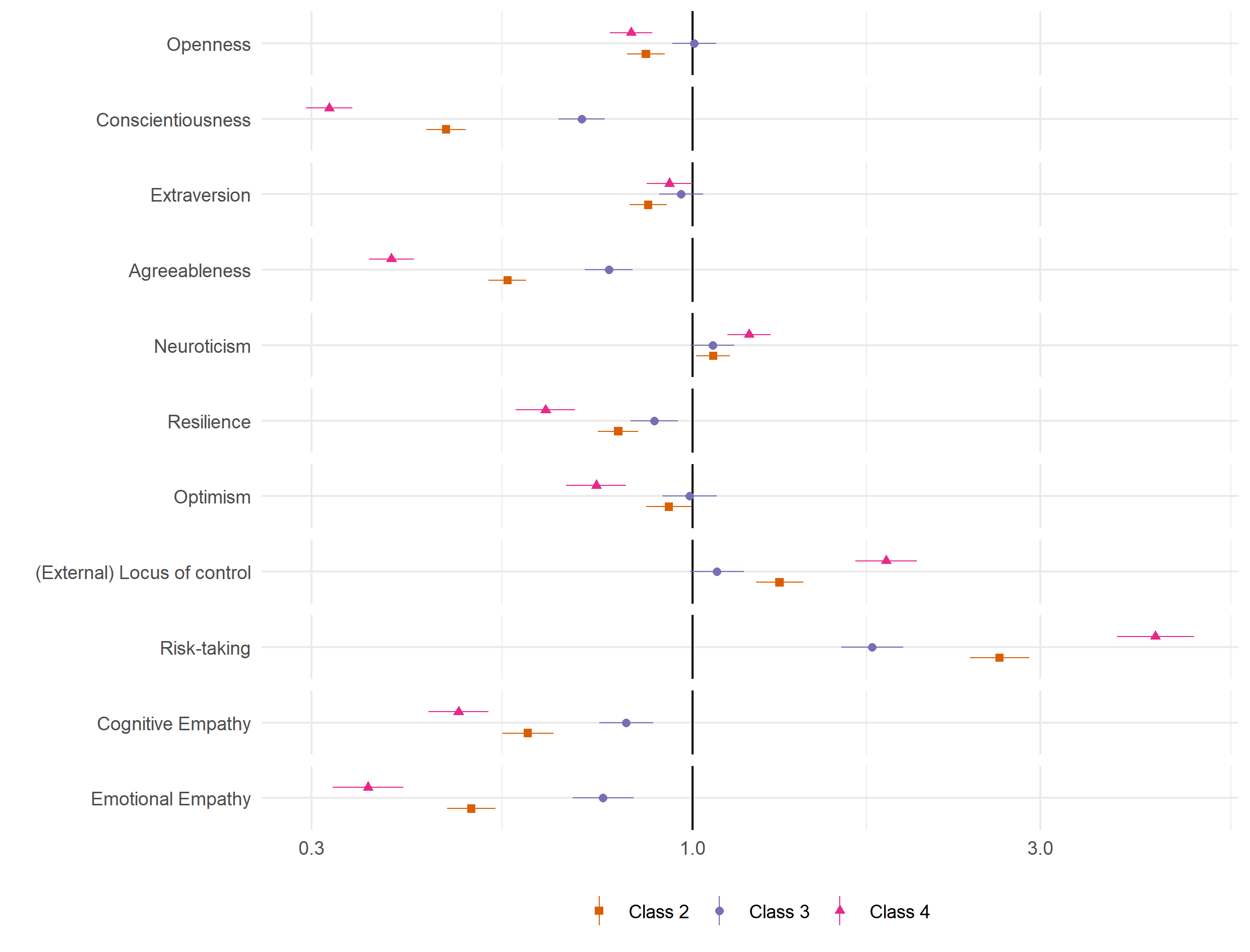

Figure S7: Result of bivariate multinomial logit models regressing (pseudo-)class membership on personality variables (reference class: Class 1). (Weighted) multiply imputed data combined using Rubin’s (1987) rules.

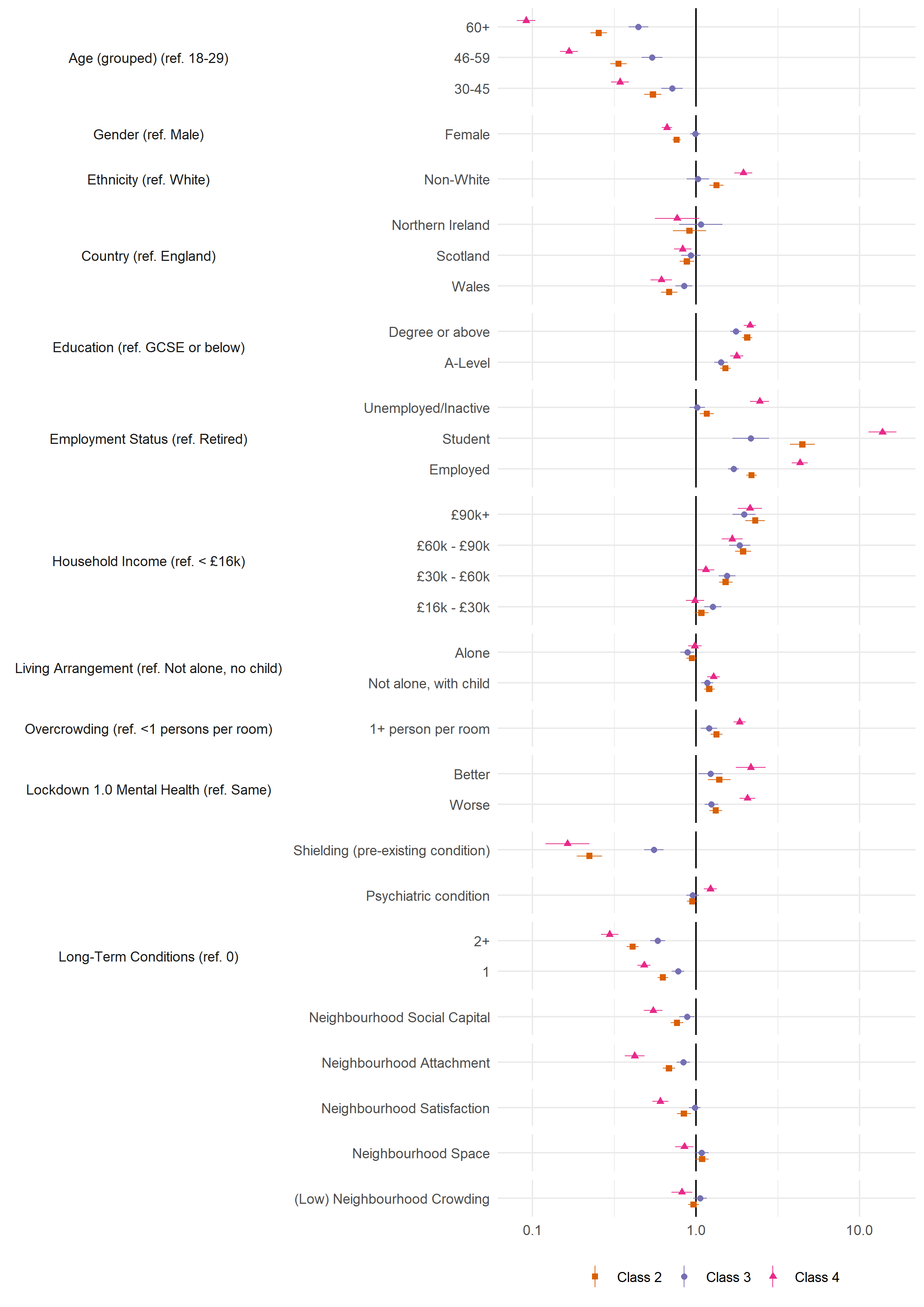

Figure S7: Result of bivariate multinomial logit models regressing (pseudo-)class membership on demographic, socioeconomic, health and neighbourhood factors (reference class: Class 1). (Weighted) multiply imputed data combined using Rubin’s (1987) rules.

### Tables

Table S1: Descriptive statistics by month of last data collection. For continuous variables, Mean (SD). For categorial variables, n (%).

|  | Variable | Ineligible | April | May | June | July | August | September | October | November | December | January | February |
| --- | --- | --- | --- | --- | --- | --- | --- | --- | --- | --- | --- | --- | --- |
|  | n | 20,613 | 3,184 | 4,209 | 2,398 | 1,532 | 3,055 | 4,095 | 2,342 | 2,029 | 1,110 | 7,785 | 19,112 |
| Age (grouped) | 18-29 | 3,911 (5.47%) | 388 (0.54%) | 590 (0.83%) | 302 (0.42%) | 185 (0.26%) | 254 (0.36%) | 420 (0.59%) | 213 (0.3%) | 169 (0.24%) | 93 (0.13%) | 334 (0.47%) | 726 (1.02%) |
|  | 30-45 | 8,571 (11.99%) | 1,304 (1.82%) | 1,641 (2.3%) | 929 (1.3%) | 512 (0.72%) | 993 (1.39%) | 1,455 (2.04%) | 802 (1.12%) | 695 (0.97%) | 339 (0.47%) | 1,440 (2.02%) | 3,466 (4.85%) |
|  | 46-59 | 5,390 (7.54%) | 983 (1.38%) | 1,309 (1.83%) | 810 (1.13%) | 533 (0.75%) | 1,010 (1.41%) | 1,317 (1.84%) | 803 (1.12%) | 695 (0.97%) | 377 (0.53%) | 2,588 (3.62%) | 6,070 (8.49%) |
|  | 60+ | 2,741 (3.84%) | 509 (0.71%) | 669 (0.94%) | 357 (0.5%) | 302 (0.42%) | 798 (1.12%) | 903 (1.26%) | 524 (0.73%) | 470 (0.66%) | 301 (0.42%) | 3,423 (4.79%) | 8,850 (12.38%) |
| Gender | Male | 5,379 (7.56%) | 766 (1.08%) | 1,031 (1.45%) | 577 (0.81%) | 366 (0.51%) | 658 (0.93%) | 958 (1.35%) | 518 (0.73%) | 421 (0.59%) | 257 (0.36%) | 1,989 (2.8%) | 4,865 (6.84%) |
|  | Female | 15,113 (21.25%) | 2,403 (3.38%) | 3,163 (4.45%) | 1,809 (2.54%) | 1,156 (1.63%) | 2,385 (3.35%) | 3,113 (4.38%) | 1,812 (2.55%) | 1,591 (2.24%) | 850 (1.2%) | 5,769 (8.11%) | 14,177 (19.93%) |
| Ethnicity | White | 18,644 (26.19%) | 2,999 (4.21%) | 3,911 (5.49%) | 2,210 (3.1%) | 1,446 (2.03%) | 2,884 (4.05%) | 3,856 (5.42%) | 2,206 (3.1%) | 1,936 (2.72%) | 1,043 (1.46%) | 7,444 (10.46%) | 18,448 (25.91%) |
|  | Non-White | 1,869 (2.63%) | 174 (0.24%) | 288 (0.4%) | 170 (0.24%) | 83 (0.12%) | 160 (0.22%) | 230 (0.32%) | 129 (0.18%) | 87 (0.12%) | 62 (0.09%) | 320 (0.45%) | 600 (0.84%) |
| Country | England | 17,209 (24.08%) | 2,603 (3.64%) | 3,391 (4.75%) | 2,027 (2.84%) | 1,250 (1.75%) | 2,519 (3.52%) | 3,349 (4.69%) | 1,886 (2.64%) | 1,688 (2.36%) | 918 (1.28%) | 6,251 (8.75%) | 15,266 (21.36%) |
|  | Wales | 1,663 (2.33%) | 218 (0.31%) | 464 (0.65%) | 207 (0.29%) | 167 (0.23%) | 328 (0.46%) | 412 (0.58%) | 259 (0.36%) | 197 (0.28%) | 105 (0.15%) | 1,026 (1.44%) | 2,555 (3.58%) |
|  | Scotland | 1,440 (2.02%) | 316 (0.44%) | 291 (0.41%) | 149 (0.21%) | 103 (0.14%) | 181 (0.25%) | 287 (0.4%) | 162 (0.23%) | 130 (0.18%) | 74 (0.1%) | 431 (0.6%) | 1,115 (1.56%) |
|  | Northern Ireland | 301 (0.42%) | 47 (0.07%) | 63 (0.09%) | 15 (0.02%) | 12 (0.02%) | 27 (0.04%) | 47 (0.07%) | 35 (0.05%) | 14 (0.02%) | 13 (0.02%) | 77 (0.11%) | 176 (0.25%) |
| Education | GCSE or below | 3,649 (5.11%) | 494 (0.69%) | 578 (0.81%) | 284 (0.4%) | 190 (0.27%) | 381 (0.53%) | 491 (0.69%) | 299 (0.42%) | 249 (0.35%) | 136 (0.19%) | 1,103 (1.54%) | 2,790 (3.9%) |
|  | A-Level | 4,128 (5.78%) | 584 (0.82%) | 784 (1.1%) | 400 (0.56%) | 262 (0.37%) | 534 (0.75%) | 730 (1.02%) | 434 (0.61%) | 334 (0.47%) | 166 (0.23%) | 1,329 (1.86%) | 3,263 (4.57%) |
|  | Degree or above | 12,836 (17.96%) | 2,106 (2.95%) | 2,847 (3.98%) | 1,714 (2.4%) | 1,080 (1.51%) | 2,140 (2.99%) | 2,874 (4.02%) | 1,609 (2.25%) | 1,446 (2.02%) | 808 (1.13%) | 5,353 (7.49%) | 13,059 (18.27%) |
| Employment Status | Retired | 1,630 (2.28%) | 322 (0.45%) | 381 (0.53%) | 221 (0.31%) | 179 (0.25%) | 521 (0.73%) | 574 (0.8%) | 346 (0.48%) | 293 (0.41%) | 197 (0.28%) | 2,546 (3.56%) | 6,644 (9.3%) |
|  | Employed | 14,886 (20.83%) | 2,281 (3.19%) | 3,063 (4.29%) | 1,768 (2.47%) | 1,081 (1.51%) | 2,124 (2.97%) | 2,846 (3.98%) | 1,622 (2.27%) | 1,428 (2%) | 753 (1.05%) | 4,328 (6.06%) | 10,369 (14.51%) |
|  | Student | 1,300 (1.82%) | 153 (0.21%) | 214 (0.3%) | 126 (0.18%) | 89 (0.12%) | 117 (0.16%) | 192 (0.27%) | 102 (0.14%) | 78 (0.11%) | 44 (0.06%) | 153 (0.21%) | 331 (0.46%) |
|  | Unemployed/Inactive | 2,797 (3.91%) | 428 (0.6%) | 551 (0.77%) | 283 (0.4%) | 183 (0.26%) | 293 (0.41%) | 483 (0.68%) | 272 (0.38%) | 230 (0.32%) | 116 (0.16%) | 758 (1.06%) | 1,768 (2.47%) |
| Household Income | < £16k | 3,230 (4.98%) | 429 (0.66%) | 560 (0.86%) | 261 (0.4%) | 199 (0.31%) | 360 (0.55%) | 504 (0.78%) | 339 (0.52%) | 248 (0.38%) | 138 (0.21%) | 1,069 (1.65%) | 2,588 (3.99%) |
|  | £16k - £30k | 4,228 (6.52%) | 638 (0.98%) | 756 (1.17%) | 425 (0.66%) | 303 (0.47%) | 619 (0.95%) | 831 (1.28%) | 444 (0.68%) | 426 (0.66%) | 239 (0.37%) | 1,871 (2.88%) | 4,581 (7.06%) |
|  | £30k - £60k | 6,201 (9.56%) | 993 (1.53%) | 1,415 (2.18%) | 807 (1.24%) | 464 (0.72%) | 997 (1.54%) | 1,325 (2.04%) | 742 (1.14%) | 654 (1.01%) | 338 (0.52%) | 2,372 (3.66%) | 6,079 (9.37%) |
|  | £60k - £90k | 2,970 (4.58%) | 514 (0.79%) | 653 (1.01%) | 416 (0.64%) | 259 (0.4%) | 494 (0.76%) | 655 (1.01%) | 327 (0.5%) | 313 (0.48%) | 171 (0.26%) | 1,009 (1.56%) | 2,320 (3.58%) |
|  | £90k+ | 2,197 (3.39%) | 334 (0.51%) | 495 (0.76%) | 304 (0.47%) | 186 (0.29%) | 324 (0.5%) | 419 (0.65%) | 294 (0.45%) | 226 (0.35%) | 124 (0.19%) | 675 (1.04%) | 1,518 (2.34%) |
| Living Arrangement | Not alone, no child | 9,926 (13.89%) | 1,515 (2.12%) | 2,070 (2.9%) | 1,191 (1.67%) | 789 (1.1%) | 1,569 (2.2%) | 2,092 (2.93%) | 1,170 (1.64%) | 1,053 (1.47%) | 596 (0.83%) | 4,534 (6.34%) | 11,288 (15.8%) |
|  | Not alone, with child | 7,810 (10.93%) | 1,209 (1.69%) | 1,487 (2.08%) | 839 (1.17%) | 474 (0.66%) | 947 (1.33%) | 1,305 (1.83%) | 802 (1.12%) | 609 (0.85%) | 327 (0.46%) | 1,510 (2.11%) | 3,537 (4.95%) |
|  | Alone | 2,877 (4.03%) | 460 (0.64%) | 652 (0.91%) | 368 (0.51%) | 269 (0.38%) | 539 (0.75%) | 698 (0.98%) | 370 (0.52%) | 367 (0.51%) | 187 (0.26%) | 1,741 (2.44%) | 4,287 (6%) |
| Overcrowding | <1 persons per room | 15,760 (22.05%) | 2,654 (3.71%) | 3,511 (4.91%) | 2,029 (2.84%) | 1,337 (1.87%) | 2,707 (3.79%) | 3,540 (4.95%) | 2,068 (2.89%) | 1,768 (2.47%) | 988 (1.38%) | 7,256 (10.15%) | 17,842 (24.97%) |
|  | 1+ person per room | 4,853 (6.79%) | 530 (0.74%) | 698 (0.98%) | 369 (0.52%) | 195 (0.27%) | 348 (0.49%) | 555 (0.78%) | 274 (0.38%) | 261 (0.37%) | 122 (0.17%) | 529 (0.74%) | 1,270 (1.78%) |
| Lockdown 1.0 Mental Health | Same |  |  |  | 363 (1.25%) | 649 (2.23%) | 952 (3.27%) | 836 (2.87%) | 516 (1.77%) | 485 (1.66%) | 300 (1.03%) | 3,634 (12.47%) | 9,501 (32.59%) |
|  | Worse |  |  |  | 319 (1.09%) | 494 (1.69%) | 698 (2.39%) | 616 (2.11%) | 366 (1.26%) | 374 (1.28%) | 210 (0.72%) | 1,872 (6.42%) | 4,593 (15.75%) |
|  | Better |  |  |  | 61 (0.21%) | 110 (0.38%) | 152 (0.52%) | 136 (0.47%) | 78 (0.27%) | 90 (0.31%) | 45 (0.15%) | 458 (1.57%) | 1,147 (3.93%) |
| Shielding (pre-existing condition) | No | 6,330 (12.02%) | 2,996 (5.69%) | 3,866 (7.34%) | 1,935 (3.67%) | 1,239 (2.35%) | 2,524 (4.79%) | 3,355 (6.37%) | 1,804 (3.43%) | 1,546 (2.94%) | 874 (1.66%) | 6,494 (12.33%) | 16,040 (30.46%) |
|  | Yes | 406 (0.77%) | 188 (0.36%) | 235 (0.45%) | 110 (0.21%) | 91 (0.17%) | 166 (0.32%) | 250 (0.47%) | 138 (0.26%) | 117 (0.22%) | 65 (0.12%) | 535 (1.02%) | 1,362 (2.59%) |
| Psychiatric condition | No | 15,848 (22.18%) | 2,424 (3.39%) | 3,280 (4.59%) | 1,872 (2.62%) | 1,202 (1.68%) | 2,391 (3.35%) | 3,175 (4.44%) | 1,816 (2.54%) | 1,609 (2.25%) | 905 (1.27%) | 6,512 (9.11%) | 16,336 (22.86%) |
|  | Yes | 4,765 (6.67%) | 760 (1.06%) | 929 (1.3%) | 526 (0.74%) | 330 (0.46%) | 664 (0.93%) | 920 (1.29%) | 526 (0.74%) | 420 (0.59%) | 205 (0.29%) | 1,273 (1.78%) | 2,776 (3.88%) |
| Long-Term Conditions | 0 | 12,394 (18.47%) | 1,849 (2.76%) | 2,556 (3.81%) | 1,504 (2.24%) | 916 (1.37%) | 1,755 (2.62%) | 2,382 (3.55%) | 1,335 (1.99%) | 1,126 (1.68%) | 622 (0.93%) | 4,090 (6.1%) | 9,944 (14.82%) |
|  | 1 | 4,390 (6.54%) | 756 (1.13%) | 937 (1.4%) | 504 (0.75%) | 336 (0.5%) | 743 (1.11%) | 911 (1.36%) | 551 (0.82%) | 523 (0.78%) | 276 (0.41%) | 2,048 (3.05%) | 5,260 (7.84%) |
|  | 2+ | 2,241 (3.34%) | 411 (0.61%) | 465 (0.69%) | 233 (0.35%) | 193 (0.29%) | 384 (0.57%) | 563 (0.84%) | 313 (0.47%) | 253 (0.38%) | 143 (0.21%) | 1,214 (1.81%) | 2,969 (4.43%) |
|  | Openness | 15.37 (3.51) | 15.41 (3.34) | 15.43 (3.46) | 15.57 (3.29) | 15.52 (3.28) | 15.5 (3.31) | 15.58 (3.32) | 15.49 (3.27) | 15.52 (3.35) | 15.64 (3.24) | 15.32 (3.28) | 15.3 (3.24) |
|  | Conscientiousness | 15.64 (3.13) | 15.6 (3.12) | 15.77 (3.04) | 15.75 (3.09) | 16 (3.03) | 15.9 (3.01) | 15.73 (3.02) | 15.75 (3.03) | 15.93 (2.94) | 15.87 (2.9) | 16.01 (2.92) | 16.08 (2.89) |
|  | Extraversion | 13.29 (4.24) | 13.21 (4.35) | 13.01 (4.39) | 13.05 (4.24) | 13.03 (4.33) | 13.01 (4.27) | 12.99 (4.27) | 12.87 (4.31) | 12.95 (4.31) | 13.02 (4.32) | 12.77 (4.3) | 12.82 (4.25) |
|  | Agreeableness | 15.67 (3.14) | 15.61 (3.1) | 15.58 (3.15) | 15.51 (3.08) | 15.55 (3.19) | 15.63 (3.1) | 15.59 (3.08) | 15.58 (3.19) | 15.61 (2.98) | 15.46 (3.07) | 15.57 (3) | 15.54 (3.04) |
|  | Neuroticism | 12.14 (4.41) | 11.88 (4.43) | 11.91 (4.44) | 11.77 (4.22) | 11.53 (4.36) | 11.62 (4.34) | 11.74 (4.39) | 11.7 (4.35) | 11.63 (4.3) | 11.45 (4.29) | 11.08 (4.25) | 10.9 (4.24) |
|  | Resilience |  |  | 19.52 (5.21) | 19.85 (5.18) | 20.05 (5.22) | 19.82 (5.21) | 19.57 (5.27) | 19.89 (5.34) | 20.06 (5.22) | 20.14 (5.22) | 20.31 (5.1) | 20.48 (5.12) |
|  | Optimism |  |  |  | 19.17 (4.84) | 19.42 (4.63) | 19.52 (4.77) | 19.29 (4.93) | 19.29 (4.78) | 19.46 (4.79) | 19.45 (4.63) | 19.88 (4.61) | 19.97 (4.65) |
|  | (External) Locus of control |  |  |  | 12.46 (2.67) | 12.35 (2.74) | 12.36 (2.64) | 12.44 (2.8) | 12.51 (2.75) | 12.32 (2.65) | 12.33 (2.61) | 12.2 (2.59) | 12.19 (2.61) |
|  | Risk-taking |  |  |  |  |  | 4.56 (2.39) | 4.51 (2.29) | 4.47 (2.33) | 4.56 (2.32) | 4.22 (2.3) | 4.34 (2.34) | 4.37 (2.35) |
|  | Cognitive Empathy |  |  |  | 18.61 (4.68) | 18.93 (4.95) | 18.8 (4.99) | 18.95 (4.84) | 18.64 (4.75) | 19.06 (4.73) | 18.96 (4.99) | 18.67 (4.8) | 18.68 (4.82) |
|  | Emotional Empathy |  |  |  | 20.9 (4.6) | 20.92 (4.69) | 20.99 (4.77) | 21.07 (4.72) | 21.08 (4.57) | 21.38 (4.49) | 21.03 (4.62) | 20.66 (4.62) | 20.66 (4.63) |
|  | Neighbourhood Social Capital |  |  |  |  | 16.31 (3.59) | 16.84 (3.6) | 16.49 (3.55) | 16.72 (3.67) | 16.8 (3.58) | 16.66 (3.53) | 17 (3.44) | 17.04 (3.46) |
|  | Neighbourhood Attachment |  |  |  |  | 10.14 (3.36) | 10.64 (3.33) | 10.26 (3.4) | 10.42 (3.34) | 10.68 (3.3) | 10.62 (3.32) | 10.96 (3.23) | 10.99 (3.2) |
|  | Neighbourhood Satisfaction |  |  |  |  | 3.95 (0.99) | 4.03 (0.95) | 3.96 (0.99) | 4.01 (0.96) | 4.05 (0.95) | 4.01 (0.95) | 4.11 (0.92) | 4.11 (0.91) |
|  | Neighbourhood Space |  |  |  |  | 8.25 (1.29) | 8.39 (1.16) | 8.3 (1.24) | 8.35 (1.24) | 8.36 (1.27) | 8.38 (1.16) | 8.42 (1.14) | 8.42 (1.12) |
|  | (Low) Neighbourhood Crowding |  |  |  |  | 6.84 (1.95) | 6.9 (1.91) | 6.95 (1.88) | 6.98 (1.87) | 6.81 (1.95) | 6.99 (1.88) | 6.98 (1.86) | 6.99 (1.86) |

### References

Davis, M. H. (1983). Measuring individual differences in empathy: Evidence for a multidimensional approach. *Journal of Personality and Social Psychology*, *44*(1), 113–126. https://doi.org/10.1037/0022-3514.44.1.113

Dohmen, T., Falk, A., Huffman, D., Sunde, U., Schupp, J., & Wagner, G. G. (2011). INDIVIDUAL RISK ATTITUDES: MEASUREMENT, DETERMINANTS, AND BEHAVIORAL CONSEQUENCES. *Journal of the European Economic Association*, *9*(3), 522–550. https://doi.org/10.1111/j.1542-4774.2011.01015.x

Hale, T., Angrist, N., Cameron-Blake, E., Hallas, L., Kira, B., Majumdar, S., Petherick, A., Phillips, T., Tatlow, H., & Webster, S. (2020). *Oxford COVID-19 Government Response Tracker*. Blavatnik School of Government. https://www.bsg.ox.ac.uk/research/research-projects/coronavirus-government-response-tracker

Hirschman, C., & Almgren, G. (2012). *University of Washington - Beyond High School (UW-BHS): Version 5* [Data set]. ICPSR - Interuniversity Consortium for Political and Social Research. https://doi.org/10.3886/ICPSR33321.V5

Mujahid, M. S., Diez Roux, A. V., Morenoff, J. D., & Raghunathan, T. (2007). Assessing the Measurement Properties of Neighborhood Scales: From Psychometrics to Ecometrics. *American Journal of Epidemiology*, *165*(8), 858–867. https://doi.org/10.1093/aje/kwm040

Rubin, D. B. (1987). *Multiple Imputation for Nonresponse in Surveys*. John Wiley & Sons, Ltd. https://doi.org/10.1002/9780470316696

Scheier, M. F., Carver, C. S., & Bridges, M. W. (1994). Distinguishing optimism from neuroticism (and trait anxiety, self-mastery, and self-esteem): A reevaluation of the Life Orientation Test. *Journal of Personality and Social Psychology*, *67*(6), 1063–1078. https://doi.org/10.1037/0022-3514.67.6.1063

Smith, B. W., Dalen, J., Wiggins, K., Tooley, E., Christopher, P., & Bernard, J. (2008). The brief resilience scale: Assessing the ability to bounce back. *International Journal of Behavioral Medicine*, *15*(3), 194–200. https://doi.org/10.1080/10705500802222972

Soto, C. J., & John, O. P. (2017). The next Big Five Inventory (BFI-2): Developing and assessing a hierarchical model with 15 facets to enhance bandwidth, fidelity, and predictive power. *Journal of Personality and Social Psychology*, *113*(1), 117–143. https://doi.org/10.1037/pspp0000096
